# Unraveling the immune signature of herpes zoster: Insights into pathophysiology and the HLA risk profile

**DOI:** 10.1101/2023.02.24.23286421

**Authors:** Romi Vandoren, Marlies Boeren, Jolien Schippers, Esther Bartholomeus, Nele Michels, Olivier Aerts, Julie Leysen, An Bervoets, Julien Lambert, Elke Leuridan, Johan Wens, Karin Peeters, Marie-Paule Emonds, Hilde Jansens, Arvid Suls, Viggo Van Tendeloo, Peter Ponsaerts, Peter Delputte, Benson Ogunjimi, Pieter Meysman, Kris Laukens

## Abstract

The varicella-zoster virus (VZV) infects over 95% of the population and establishes latency afterwards. Reactivation of VZV causes herpes zoster (HZ), commonly known as shingles, which presents as a painful rash in mostly the elderly and people with a weakened immune system. However, HZ might occur in otherwise healthy individuals too. In this study, we have studied the immune signature of HZ to better understand HZ’s pathophysiology. We provide a general overview of the antiviral state and the activation of innate and adaptive immune responses during HZ. Differential gene expression and gene ontology analyses revealed upregulation of several genes and host immune pathways during herpes zoster, especially related to type I IFN response but also related to adaptive immune responses. Intriguingly, no differences in gene expression were noted during convalescence between HZ patients and controls. Furthermore, we conducted the largest HLA association study on HZ to date using the UK Biobank and identified seven protective and four risk HLA alleles associated with the development of herpes zoster. These findings reveal key genes and pathways involved in the host immune response to symptomatic VZV reactivation and provide new molecular insights into the development of HZ.

## INTRODUCTION

Herpes zoster (HZ, shingles) is caused by symptomatic reactivation of the varicella-zoster virus (VZV) and typically presents as a painful dermatomal rash, frequently accompanied by mild symptoms such as fever, headache, and fatigue [1]. Post-herpetic neuralgia (PHN), i.e. long-lasting neuropathic pain, is the most common complication of HZ and extensively adds to the burden of disease [2, 3]. Naturally, over 95% of the population gets infected with VZV, resulting in chickenpox (varicella) [1, 4]. Upon resolution of a primary infection with VZV (varicella), VZV particles gain access to neural ganglia where latency is established [1]. At a later age, VZV can reactivate to cause HZ [1].

The lifetime risk of developing HZ is between 25% and 30%, rising to 50% in those aged at least 80 years [3, 5-7]. Indeed, increasing age is a well-known risk factor for the development of HZ, owing to a decline in cellular immunity, i.e. immunosenescence [8, 9]. Yet, HZ also frequently occurs in immunocompromised patients [10] and otherwise healthy individuals too [3, 11]. Besides waning cellular immunity due to aging, the constitution of the T cell receptor (TCR) repertoire could also be a determining factor for the development of HZ. During the development of T cells in an individual, the TCR repertoire will reflect the major histocompatibility complex (MHC) constitution of the individual [12]. Thus, depending on an individual’s MHC constitution encoded by HLA genes, each individual will be able to recognize a different set of (virus-derived) peptides and will generate a different immune response [12]. This is a major factor in the individuality of the immune response [12]. As such an individual’s HLA genetic constitution could be a risk factor for the development of HZ [12]. The nine so-called classical MHC genes are HLA-A, -B, -C belonging to MHC class I, and HLA-DPA1, -DPB1, -DQA1, -DQB1, HLA-DRA, and HLA-DRB1 belonging to MHC class II [12]. A previous pilot study including 50 Belgian individuals with a history of HZ and a control population of 25,000 Belgians obtained via the Red Cross, already showed that HLA-A*11 was protective, whereas HLA-B*37 was a risk allele for the development of HZ [13]. In addition, a meta-analysis showed that HLA-A*02 and HLA-B*40 were protective, whereas HLA-A*33 and HLA-B*44 were risk alleles for PHN in Japanese patients [14]. To further investigate HLA genetic constitution as a risk factor for the development of HZ, we carried out an HLA-association study on data from the UK biobank.

Besides HLA genetic variation, other genetic polymorphisms, especially related to immune system genes, can impact an individual’s likeliness to develop HZ. Following VZV infection, VZV is immediately sensed by the innate immune system via pattern-recognition receptors (PRRs) like RNA Polymerase III [15-19]. Subsequent activation of downstream pathways leads to the production of type I IFNs and proinflammatory cytokines that inhibit viral replication and recruit inflammatory cells to the site of infection [16, 20]. Binding of these type I IFNs to their receptor ultimately leads to the induction of a large range of interferon-stimulated genes (ISGs) with direct antiviral effector functions [21]. The importance of an adequate type I IFN response to control VZV infection is illustrated by several studies reporting that mutations in *POLR3, TLR3, STAT1, STAT2, TYK2* and *NEMO* lead to increased susceptibility to VZV infection or even VZV viral encephalitis [18, 22-27]. In addition to type I IFN direct antiviral effector functions, they also negatively regulate IL-12 expression, activate NK cell-mediated lysis, and promote T cell expansion and activation [15]. Besides type I IFN, more recently type III IFN (IFN-λ) was shown to exert direct antiviral activity and promote T cell Th1 phenotype, similar to a type I IFN response [28, 29]. Finally, the production of type II IFNs (IFN-y) also results in the production of ISGs. IFN-y production is, however, tightly controlled and restricted to specialized immune cells, predominantly CD4+ and CD8+ T cells and NK cells [30, 31]. As a second line of defense, VZV-specific antibodies and VZV-specific cellular immunity, is activated [32]. CD4+ and CD8+ T effector and memory cells are essential for recovery and memory T cells that develop during a primary infection are necessary to prevent VZV reactivation [32].

Differential gene expression (DGE) and gene ontology (GO) analyses have been widely used to provide insights into the pathophysiology of (viral) diseases. They can also lead to identification of novel biomarkers and provide clues for the development of therapeutics or vaccines. Since these analyses show genes and pathways that are significantly up- or downregulated during a disease, they also give a view of genes and/or pathways that might be risk factors for the development of these diseases. In this study we performed DGE and GO analyses on blood transcriptome data from HZ patients during HZ and convalescence.

## MATERIALS AND METHODS

### Participants

A total cohort of twenty-six herpes zoster patients aged between 18 years and 70 years (median age 51 years; 13 men, 13 women) were prospectively recruited during an active HZ episode, as confirmed by a positive VZV PCR on skin swab, saliva or blood (EDTA) or by significantly elevated (> 4 times) VZV IgG titers. The exclusion criteria were: (i) auto-immune disease, immunosuppressed state or immunocompromised state due to disease or medication (e.g. use of systemic prednisolone or equivalent > 20mg/d for more than two weeks in the last six months; intra-articular, inhalation and topical steroids are allowed), (ii) invasive malignancy <20y prior to the study, (iii) serious disorders of coagulation, (iv) any condition that the investigator believed might interfere with the study. During the first blood collection a PAXgene RNA tube was taken from each HZ patient and kept at RT for at least 2h before moving to -80°C. A second PAXgene RNA tube was collected one year after the HZ episode (23 out of the initial 26 HZ participants: 11 men, 12 women). Age (± 1 birth year) and sex matched controls without a history of HZ were recruited for each HZ patient and donated blood around the one-year timepoint. To optimize the inclusion of controls, PAXgene RNA of three of the controls were collected during sampling via another study with approval by the IRB. Six of the controls also donated a PAXgene RNA tube one year earlier around the same time as blood collection from their matched HZ patients during the active HZ episode, which served as an internal quality control for storage and handling. This study was approved by the ethics board of the Antwerp University Hospital and University of Antwerp. All methods were performed in accordance with the relevant guidelines and regulations when applicable. Written informed consent was obtained from all study participants.

### RNA extraction

RNA extraction from whole blood collected in PAXGene tubes was performed via a column-based RNA extraction using the PaxGene blood RNA extraction kit (Cat. No./ID: 762174, Qiagen) following the manufacturer’s instructions. Subsequently, RNA was concentrated using the RNA clean & concentrator-5 kit (R1013, Zymo research). RNA concentration was measured using the Qubit™ RNA BR Assay Kit (Cat n°: Q10211, Thermo Fisher) on a Qubit 4 fluorometer (Invitrogen).

### 3’ mRNA sequencing

All RNA samples were prepared with the QuantSeq 3’ mRNA-Seq Library Prep Kit FWD for Illumina (Lexogen GmbH) following the standard protocol for long fragments. Resulting cDNA libraries were equimolarly pooled, up to 40 samples for one NextSeq 500/550 sequencing run (high output v2.5 kit, 150 cycles, single read, Illumina). RNA isolated from PAXGene tubes, collected from the same individual (during HZ and one year after HZ onset) and their age and sex matched controls, was sequenced in the same run to avoid inter-assay variability. This resulted in on average 10.5 million reads for each sample after quality filtering.

### NGS data processing

Raw data from the NextSeq was demultiplexed and further processed through an in-house developed 3’mRNA sequencing pipeline. The quality of all reads was evaluated using FastQC (v0.11.5) before and after processing with Trimmomatic (v0.36). Trimmomatic was used to remove the leading 20 bases from reads, ensure a minimum quality score of 15 over a sliding window of 4 bases and require a minimum read length of 30 bases. The use of oligodT primers can potentially cause poly-A stretches at the 3’ end. To remove these poly-A stretches, the 3’ read end was trimmed with an in-house developed poly-A removal script. All remaining sequences were mapped against the Ensembl human reference genome build 38 with HISAT2 (v2.0.4). HTseq (v0.6.1) was used to count all reads for each gene and set up a read count table.

### Differential gene expression analysis and gene ontology enrichment analysis

Differential gene expression analysis was performed using the DESeq2 Bioconductor package [33]. Gene ontology (GO) enrichment analysis was performed on significantly differentially expressed genes using topGO [34]. To determine significantly enriched/depleted gene ontology terms related to biological processes, a Fisher’s exact test was performed with Benjamini-Hochberg correction for multiple testing (FDR < 0.05). As a reference background set of genes, we used all genes (total number: 35894) measured during the 3’-mRNAseq experiment. All codes used for the pre-processing and analysis of the data within this manuscript as well as the codes used to generate results and the DESeq2 results itself, are publicly available on github at github.com/pmeysman/VZV-RNAseq. Heat map clustering was performed using the pheatmap R package v 1.0.12.

### UK biobank data acquisition

The data for the HLA association analysis was obtained from the population level exome OQFE variants (200k release) in the UK Biobank [35]. Whole-exome sequencing data in the UK Biobank was generated using the IDT xGen Exome Research Panel v1.0 including supplemental probes. Multiplexed samples were sequenced with dual-indexed 75 × 75 bp paired-end reads on the Illumina NovaSeq 6000 platform using S2 and S4 flow cells. The study population for this project was selected based on specific inclusion and exclusion criteria relating to risk factors for HZ development such as age, asthma, chronic obstructive pulmonary disease, depression, malignancy, other disorders, and those taking biologicals or corticosteroids (Supplementary Table S1). This ultimately resulted in 2830 individuals diagnosed with herpes zoster and 135,992 healthy control individuals from the UK biobank data. For these selected individuals, unique HLA alleles were collected from the UK biobank.

### HLA association study

To investigate the potential association between HLA background and susceptibility to herpes zoster, an HLA association analysis was conducted. HLA frequencies for HZ-afflicted patients (n=2830) and controls (n=135992) were extracted from the UK Biobank at the level of unique HLA alleles (e.g. HLA-A*01:01). Here, the control group is defined as UK Biobank participants that did not have a history of HZ at the time of sampling. For each of the patient and control groups, the HLA allele frequency counts were summed over all patients in that group. This resulted in a confusion matrix for each unique HLA allele, which describes the frequency of a certain HLA allele in herpes zoster patients compared to healthy controls. Next, a Fisher’s exact test was used to calculate the P value and odds ratio for each allele to identify enriched or depleted HLA alleles in the patient group. Benjamini-Hochberg multiple testing correction was applied to limit the false discovery rate and significant associations (P value≤0.05) were identified.

As HLA background can vary significantly between different ethnicities in the data, an additional analysis was performed where the data was split based on population values obtained from the UK Biobank. This resulted in three groups depicting a ‘White’, ‘Black’ or ‘Asian’ background, conform to the labeling used in the UK biobank. The same HLA association workflow and statistical testing was then applied to each of these subsets in the data.

## RESULTS

### Differential gene expression analysis

We first compared the gene expression profile from blood of HZ patients taken during the HZ episode with those one year after the HZ episode and found 841 DEGs (596 upregulated and 245 downregulated genes: Supplementary Table S2). The top 20 most upregulated DEGs during HZ compared to one year after the HZ episode are shown in Table 1. Seven of the top 20 upregulated genes, *IFI44, IFI44L, IFI27, RSAD2, ISG15, SERPING1* and *SIGLEC1*, are interferon-induced genes (ISGs) produced by the innate immune system upon virus encounter. In addition, *BATF2*, a transcription factor that participates in the differentiation of CD8+ thymic conventional dendritic cells following infection, was also significantly upregulated during the acute HZ episode. Furthermore, two genes associated with the antibody response: *MZB1* which associates with IgM, and *IGHG4* which codes for the constant region of immunoglobulin heavy chains, were significantly upregulated too. Other genes in the top 20 list of DEGs are not directly related to viral processes or immune responses. Noteworthy, although they did not appear in the top 20 list of DEGs, several other ISGs: *Mx1, IFIT5, OASL, IFI35, IFI27* and *STAT2*, were significantly upregulated during HZ (Supplementary Table S1). Also, RNA polymerase III (*POLR3*) subunit D (*POLR3D*) and GL (*POLR3GL*), were significantly upregulated. Finally, *CXC3* which is primarily expressed on activated T lymphocytes and NK cells was also significantly upregulated during HZ (Supplementary Table S1). No notable downregulated genes related to viral or immunological functions were found. A heat map shows clustering of gene expression profiles from blood taken during the acute HZ episode (HZ_XX, light blue) and clustering of those one year after HZ (HZ_XX.1Y, pink/red) (Figure 1).

**Table 1:** Top 20 most upregulated DEGs during HZ compared to one year after HZ.

| Gene symbol | Description | log2 Fold Change | Adj. P value |
| --- | --- | --- | --- |
| **IGHGP** | immunoglobulin heavy constant gamma P (non-functional) | 2,97 | 2,47E-08 |
| **IFI44L** | interferon induced protein 44 like | 2,42 | 1,82E-04 |
| **IFI27** | interferon alpha inducible protein 27 | 2,32 | 8,40E-04 |
| **ANKRD22** | ankyrin repeat domain 22 | 2,16 | 8,57E-04 |
| **BATF2** | basic leucine zipper ATF-like transcription factor 2 | 2,14 | 6,08E-04 |
| **RSAD2** | radical S-adenosyl methionine domain containing 2 | 2,04 | 9,71E-04 |
| **SERPING1** | serpin family G member 1 | 1,98 | 8,02E-04 |
| **PTTG1** | PTTG1 regulator of sister chromatid separation | 1,96 | 9,37E-12 |
| **ISG15** | ISG15 ubiquitin like modifier | 1,94 | 1,28E-03 |
| **ETV7-AS1** | ETV7 and PTX1 antisense RNA 1 | 1,89 | 2,24E-02 |
| **SIGLEC1** | sialic acid binding Ig like lectin 1 | 1,73 | 4,71E-03 |
| **IGHG4** | immunoglobulin heavy constant gamma 4 (G4m marker) | 1,71 | 2,36E-03 |
| **MT-TV** | mitochondrially encoded tRNA-Val (GUN) | 1,70 | 2,45E-03 |
| **CMPK2** | cytidine/uridine monophosphate kinase 2 | 1,66 | 7,52E-03 |
| **MZB1** | marginal zone B and B1 cell specific protein | 1,62 | 7,35E-05 |
| **BIRC5** | baculoviral IAP repeat containing 5 | 1,60 | 3,31E-05 |
| **FABP5** | fatty acid binding protein 5 | 1,53 | 1,65E-05 |
| **IFI44** | interferon induced protein 44 | 1,52 | 8,39E-03 |
| **FAM111B** | FAM111 trypsin like peptidase B | 1,43 | 1,38E-05 |
| **TOP2A** | DNA topoisomerase II alpha | 1,43 | 4,30E-12 |

**Figure 1:**
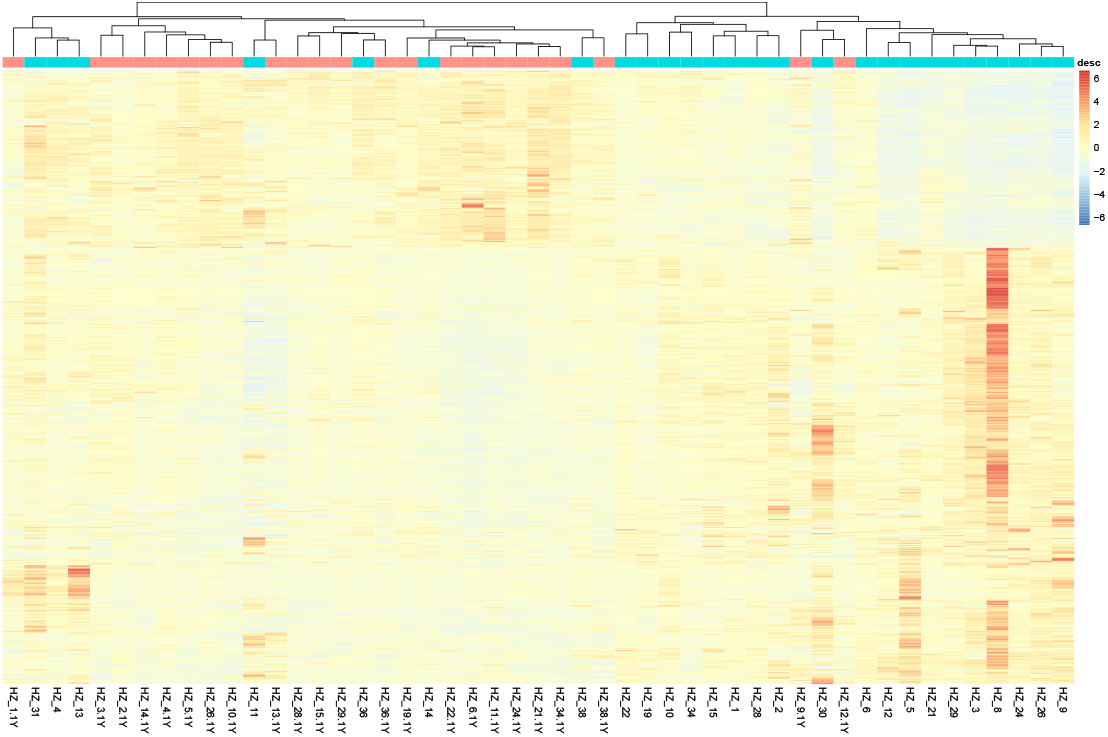
Heat map of DEGs in blood from patients taken during HZ compared to one year after HZ. Heat map shows clustering of gene expression profiles from blood taken during the acute HZ episode (HZ_XX, light blue) and clustering of those one year after the HZ episode (HZ_XX.1Y, pink/red).

Next, we compared the gene expression profiles of HZ patients during the active HZ episode with those of control participants and found 485 DEGs (341 upregulated and 144 downregulated genes: Supplementary Table S3). The top 20 most upregulated DEGs during HZ compared to control participants are shown in Table 2. Six of these genes including *IFI27, IFI44L* and *IGHG4*, were also found in the top upregulated genes when comparing acute HZ versus one year after HZ. Interestingly, *C4BPA*, an inhibitor of the classical and lectin pathways of the complement system, was upregulated by 2 log folds.

**Table 2:** Top 20 most upregulated DEGs during HZ compared to control participants.

| Gene symbol | Description | log2 Fold Change | Adj. P value |
| --- | --- | --- | --- |
| **IFI27** | interferon alpha inducible protein 27 | 2,62 | 5,82E-05 |
| **IGHGP** | immunoglobulin heavy constant gamma P (non-functional) | 2,36 | 1,63E-05 |
| **ZWINT** | ZW10 interacting kinetochore protein | 2,08 | 1,80E-05 |
| **C4BPA** | complement component 4 binding protein alpha | 2,01 | 2,05E-03 |
| **CDK1** | cyclin dependent kinase 1 | 1,81 | 3,34E-04 |
| **FAM111B** | FAM111 trypsin like peptidase B | 1,63 | 5,61E-08 |
| **IFI44L** | interferon induced protein 44 like | 1,60 | 3,08E-02 |
| **IGHG4** | immunoglobulin heavy constant gamma 4 (G4m marker) | 1,60 | 5,86E-03 |
| **BIRC5** | baculoviral IAP repeat containing 5 | 1,54 | 5,02E-05 |
| **MT-TV** | mitochondrially encoded tRNA-Val (GUN) | 1,50 | 1,07E-02 |
| **H2AC14** | H2A clustered histone 14 | 1,50 | 4,31E-03 |
| **HJURP** | Holliday junction recognition protein | 1,45 | 6,70E-04 |
| **FABP5** | fatty acid binding protein 5 | 1,40 | 5,82E-05 |
| **IGHV1OR15-1** | immunoglobulin heavy variable 1/OR15-1 (non-functional) | 1,38 | 5,86E-03 |
| **RPL9** | ribosomal protein L9 | 1,34 | 1,70E-04 |
| **PTTG1** | PTTG1 regulator of sister chromatid separation | 1,34 | 1,23E-05 |
| **TOP2A** | DNA topoisomerase II alpha | 1,33 | 7,93E-11 |
| **IGHV1OR15-1** | novel gene identicle to IGHV1OR15-1 | 1,33 | 7,05E-03 |
| **PDILT** | protein disulfide isomerase like | 1,25 | 8,23E-03 |
| **S100B** | S100 calcium binding protein B | 1,23 | 2,11E-02 |

Finally and surprisingly, when the gene expression profiles of blood from HZ patients during convalescence (one year after the HZ episode) were compared with those of control participants no DEGs were found, thereby indicating that no fundamental differences in whole blood gene expression existed between HZ patients during convalescence (one year after their episode) and controls.

In addition, as a control, we performed DGE analyses on samples from control participants taken one year apart from each other. Six DEGs were found which were all related to ribosomal processes and are unlikely to reflect biologically significant differences.

### Gene ontology analysis

When we compared gene expression profiles from zoster patients during HZ with those one year after HZ, we found 843 DEGs. GO enrichment analysis of these DEGs revealed the involvement of 290 significant GO categories (top 200, supplementary table S4) including several pathways related to viral processes and host immune responses, also those specific to viral infection. Table 3 shows the first 20 significant enriched GO categories that were related to viral processes and host immune responses. These data clearly indicate that viral processes were ongoing and that host immunity to viral infection was activated. Interestingly, six of these categories also came up in the GO enrichment analysis of the DEGs during HZ onset versus those of control participants (Table 4, Supplementary Table S5). Not surprisingly, viral transcription is one of the top GO terms in both analyses.

**Table 3:** **First 20 significant GO categories related to viral processes and host immune responses of DEGs during HZ episode versus those one year after HZ**.

| GO.ID | Term | P value |
| --- | --- | --- |
| GO:0019083 | viral transcription | 1,00E-20 |
| GO:0002479 | antigen processing and presentation of exogenous peptide antigen via MHC class I, TAP-dependent | 0,00014 |
| GO:0050852 | T cell receptor signaling pathway | 0,0002 |
| GO:0050853 | B cell receptor signaling pathway | 0,00029 |
| GO:0035722 | interleukin-12-mediated signaling pathway | 0,00042 |
| GO:0070498 | interleukin-1-mediated signaling pathway | 0,00056 |
| GO:0060337 | type I interferon signaling pathway | 0,00094 |
| GO:0051607 | defense response to virus | 0,00095 |
| GO:0016032 | viral process | 0,00102 |
| GO:0032020 | ISG15-protein conjugation | 0,00165 |
| GO:0038096 | Fc-gamma receptor signaling pathway involved in phagocytosis | 0,00459 |
| GO:0006955 | immune response | 0,01661 |
| GO:0045059 | positive thymic T cell selection | 0,01864 |
| GO:0006910 | phagocytosis, recognition | 0,02094 |
| GO:0019886 | antigen processing and presentation of exogenous peptide antigen via MHC class II | 0,02287 |
| GO:0006958 | complement activation, classical pathway | 0,02994 |
| GO:0002230 | positive regulation of defense response to virus by host | 0,0316 |
| GO:0046596 | regulation of viral entry into host cell | 0,03664 |
| GO:0019064 | fusion of virus membrane with host plasmamembrane | 0,03672 |
| GO:0050871 | positive regulation of B cell activation | 0,03989 |

**Table 4:** GO categories related to viral processes and host immune responses during HZ versus control participants.

| GO.ID | Term | P value |
| --- | --- | --- |
| GO:0019083 | viral transcription | 7,10E-15 |
| GO:0035722 | interleukin-12-mediated signaling pathway | 0,01785 |
| GO:0016032 | viral process | 0,02103 |
| GO:0002479 | antigen processing and presentation of exogenous peptide antigen via MHC class I, TAP-dependent | 0,02176 |
| GO:0006910 | phagocytosis, recognition | 0,02291 |
| GO:0070498 | interleukin-1-mediated signaling pathway | 0,03111 |

**Table 5:**
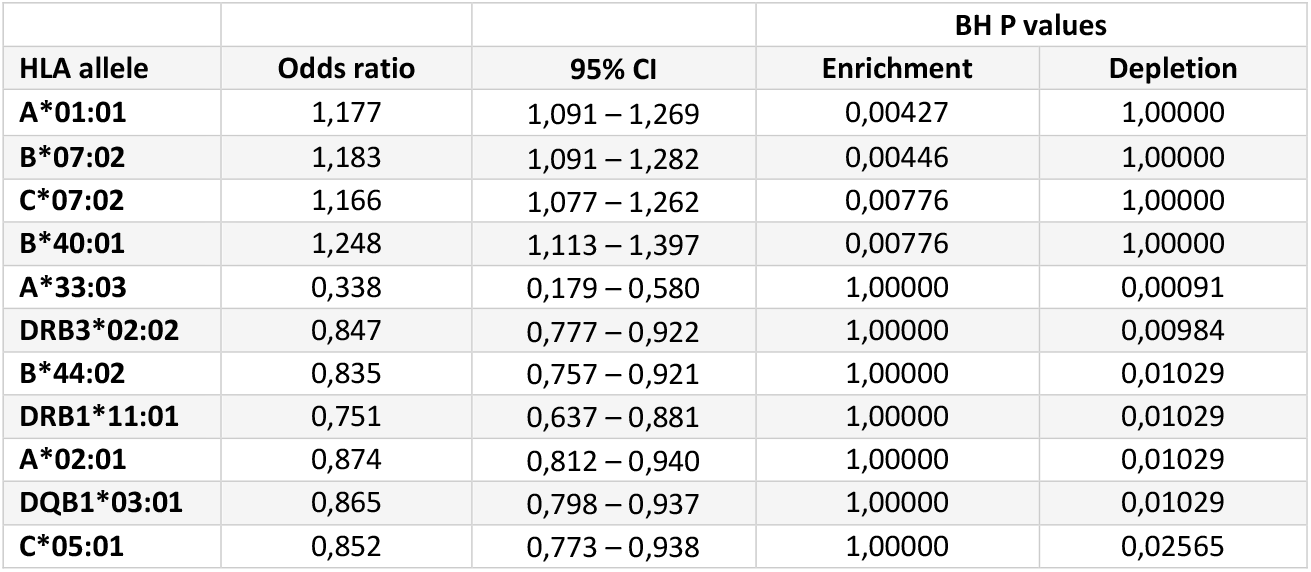
**All 11 significantly enriched or depleted HLA alleles in the complete UK Biobank dataset related to development of HZ in patients versus controls**.

### HLA association analysis

We compared HLA allele frequencies between HZ patients (n=2830) and healthy controls (n=135992) from the UK Biobank. This HLA association study revealed significant differences in HLA allele distribution between HZ patients and controls. We discovered four risk HLA alleles that were significantly enriched (P value ≤ 0.05) in HZ patients, namely HLA-A*01:01, HLA-B*07:02, HLA-C*07:02 and HLA-B*40:01. Additionally, seven protective HLA alleles were found to be significantly depleted (P value ≤ 0.05) in HZ patients, namely HLA-A*33:03, HLA-DRB3*02:02, HLA-B*44:02, HLA-DRB1*11:01, HLA-A*02:01, HLA-DQB1*03:01 and HLA-C*05:01. To account for HLA frequency differences on population level, the dataset was divided into three different groups including a ‘White’, ‘Asian’ and ‘Black’ background. Application of the same HLA association workflow on the three population subsets revealed different results for the protective and risk alleles in each subset (Supplementary Table S6). Only four unique HLA alleles were depleted in all three different cohorts, including HLA-A*33:03, DRB3*02:02, DRB1*11:01 and A*02:01 while none were enriched in all three cohorts. However, the ‘Asian’ and ‘Black’ populations only have a small fraction of HZ patients compared to control individuals (1.51% and 1.14% HZ patients respectively) resulting in a lack of statistical power to confirm these results.

## DISCUSSION

Gene expression profile analysis of blood from HZ patients during the acute HZ episode versus those one year after the HZ episode, revealed 843 DEGs. Seven of the top twenty upregulated genes were ISGs: *IFI44, IFI44L, IFI27, ISG15, RSAD2, SERPING1* and *SIGLEC1*. Curiously, *IFI44* positively affects virus production and negatively modulates innate immune responses induced after viral infections [36]. Moreover, *IFI44L* was described to negatively modulate innate immune responses induced after virus infections [37]. Upregulation of these two genes likely reflects negative feedback loops in the host antiviral response to avoid excessive inflammation. Furthermore, *IFI27* has growth inhibitory and antiviral functions and is involved in type I IFN-induced apoptosis [38, 39]. *ISG15* is another key ISG that displays direct antiviral activity against many viruses. However, an antiviral role during VZV infection or reactivation was not yet described. Interestingly, *ISG15* targets many different genes in the IFN-pathway including *Mx1* which was also significantly upregulated during HZ compared to one year after HZ [40, 41]. Viperin (virus inhibitory protein, ER-associated, interferon-inducible), encoded by the *RSAD2* gene, is another key player of the innate immune response and is induced upon viral infection. Viperin limits both DNA and RNA viruses via multiple mechanisms [42, 43]. Finally, *SERPING1* is involved in regulation of the complement cascade and *SIGLEC1* is a macrophage-restricted adhesion molecule that mediates sialic-acid dependent binding to lymphocytes. Furthermore, we found that RNA polymerase III (*POLR3*) subunit D (*POLR3D*) and GL (*POLR3GL*), were significantly upregulated during HZ. Interestingly, mutations in *POLR3A, POLR3C, POLR3E* and *POLR3F* have been associated with susceptibility to VZV-induced encephalitis and pneumonitis [44, 45]. However, such association for *POLR3* subtypes D or GL have not been described so far. Furthermore, a recent report in which the involvement of cellular calcium disorder in the development of PHN was investigated, revealed several DEGs [46]. Voltage-gated and ligand-gated calcium channels in neurons play an important role in the regulation of Ca^2+^ influx in neurons and glia of the CNS [47]. Moreover, Ca^2+^ signals are increasingly being associated with pain and other CNS diseases [47, 48]. Interestingly, in our cohort of HZ patients without known data on PHN, several DEGs related to the GO term ‘negative regulation of cytosolic calcium ion concentration’ were downregulated one year after the HZ episode: *CAB39, CACNA1D, CAMK1D* and *CACNB4*. Conversely, only one gene related to calcium signaling (*CIB1*) was upregulated one year after HZ. Additionally, two genes related to calcium signaling (*S100B* and *CALHM6*) were upregulated during acute HZ compared to controls. Thus, our data suggest that indeed calcium signaling pathways are involved in HZ pathogenesis.

Whilst we found several DEGs during HZ compared to one year after HZ, we did not find any DEGs one year after HZ versus control participants. However, some HZ patients had a very high expression of *C4BPA* (although overall not significant) compared to controls. In addition, *C4BPA* was the fourth highest expressed gene during active HZ compared to control participants. Thus, *C4BPA* upregulation may be a potential risk factor for the development of HZ. Interestingly, C4BP is a major inhibitor of the classical and lectin pathways of the complement system, making it an attractive target for pathogens to escape complement attack [49]. Moreover, C4BP can act independently of the complement against pathogens [50, 51]. Indeed, a recent study showed that direct binding of C4BP to influenza A virus subtype H1N1 suppressed viral infection, whereas binding to H3N2 subtype promoted viral infection [51]. Whether C4BP may play a pro- or anti-viral role in VZV infection or reactivation needs further investigation.

Additionally, GO enrichment analysis revealed several categories related to viral processes and host immune responses that were enriched during HZ and matched the DGE analyses: upregulation of the type I IFN signaling pathway, ISG15-protein conjugation and complement activation. GO analysis also revealed enrichment of the interleukin-1 (IL-1) and -12 (IL-12) pathway during HZ. IL-1 promotes recruitment of inflammatory cells at the site of inflammation [52], whereas IL-12 is involved in the differentiation of naive T cells into Th1 cells, stimulates IFNy and TNF-α production by T cell and NK cells and enhances NK-and CD8+ T cells cytotoxicity [53, 54]. Related to the latter, the T cell receptor signaling pathway as well as MHC Class I and Class II antigen presentation pathways were significantly enriched during HZ. Finally, B cell receptor signaling and pathways involved in phagocytosis were also significantly enriched during HZ.

Finally, the HLA association analysis of HZ patients and controls in the UK Biobank revealed that four HLA alleles are significantly enriched in HZ patients while seven other alleles are significantly decreased. These findings suggest that the HLA background plays a role in susceptibility to herpes zoster and that certain HLA alleles can confer a protective effect against HZ, while others increase the risk of developing HZ. Some of our discovered significant HLA alleles have already been associated with the development of postherpetic neuralgia in herpes zoster patients [14, 55, 56]. This includes HLA-A*33 and HLA-B*44, which are significantly enriched for patients in this study, while HLA-A*02 and HLA-B*40 are significantly depleted. When comparing these results to our own, it becomes clear that these do not necessarily overlap. However, the patients from these previous studies were mostly from Asian descent while for our cohort the largest part of the UK biobank participants fall within the ‘White’, Caucasian, subset and thus dictate the HLA association results. Interestingly, the association results for our ‘Asian’ subset matches the previously discovered associations a lot better, although without statistical significance due to the small sample size. This indicates that there are some clear differences for risk and protective alleles between different populations and that associations should be considered for each genetic background separately. Therefore, it should be noted that these findings are limited by the sample size and population characteristics of the UK Biobank data, and additional studies are needed to confirm these results. Additionally, these findings also do not prove causality or reveal the biological mechanisms underlying these associations and thus should be interpreted with caution.

In conclusion, DGE and GO enrichment analyses revealed upregulation of several host immune pathways during HZ, especially related to the type I IFN response, but related to the adaptive immune response too. A genome-wide association study in a larger cohort could reveal if genetic variants are implicated in HZ pathogenesis. In addition, a single cell RNA sequencing approach would allow the identification of cell-type specific transcriptional changes that occur. The HLA association analysis between HZ patients and healthy controls revealed four risk and seven protective alleles that vary between different populations. This implicates that additional HLA studies on specific populations could further validate our findings and uncover population differences in risk and protective allele frequencies for herpes zoster susceptibility.

## Data Availability

All data produced in the present study are available upon reasonable request to the authors

