## Supplemental Materials for "Unraveling the immune signature of herpes zoster: Insights into pathophysiology and the HLA risk profile"

### SUPPLEMENTARY MATERIALS

***Supplementary Table S1: Detailed description of the inclusion and exclusion criteria used to select HZ patients and control individuals from the UK biobank.***

| Exclusion criterium | Description |
|---|---|
| **Age** | Development of HZ after the age of 60y |
| **Malignancy** | Malignancy development ≤ 5y pre-HZ development |
| **Depression** | Depression ≤ 1y pre- or post-HZ development |
| **Asthma** | Diagnosis of asthma |
| **COPD** | Diagnosis of chronic obstructive pulmonary disease |
| **Other disorders** | Diagnosis of inflammatory bowel disease, kidney failure, rheumatoid arthritis, psoriasis or systemic lupus erythematosus |
| **Biologicals** | Use of one or more of the following: ciclosporin, tacrolimus, mycophenolic acid, sirolimus, leflunomide, anakinra, adalimumab, azathioprine and methotrexate |
| **Corticosteroids** | Use of one or more of the following: betamethasone, dexamethasone, methylprednisolone, prednisolone, prednisone, triamcinolone, cortisone and hydrocortisone |

***Supplementary Table S2: Top 200 most upregulated DEGs during HZ compared to one year after HZ.***

| uniprot_gn_symbol | ensembl_gene_id | uniprot_gn_id | description | baseMean | log2 FoldChange | lfcSE | stat | p-value | Adj p-value |
|---|---|---|---|---|---|---|---|---|---|
| | ENSG00000253755 | | immunoglobulin heavy constant gamma P (non-functional) | 12,39 | 2,97 | 0,43 | 6,89 | 5,54E-12 | 2,47E-08 |
| IFI44L | ENSG00000137959 | Q53G44 | interferon induced protein 44 like | 273,91 | 2,42 | 0,47 | 5,15 | 2,65E-07 | 1,82E-04 |
| IFI27 | ENSG00000165949 | P40305 | interferon alpha inducible protein 27 | 75,50 | 2,32 | 0,49 | 4,73 | 2,27E-06 | 8,40E-04 |
| ANKRD22 | ENSG00000152766 | Q5VYY1 | ankyrin repeat domain 22 | 69,04 | 2,16 | 0,46 | 4,72 | 2,36E-06 | 8,57E-04 |
| BATF2 | ENSG00000168062 | Q8N1L9 | basic leucine zipper ATF-like transcription factor 2 | 43,66 | 2,14 | 0,44 | 4,86 | 1,19E-06 | 6,08E-04 |
| RSAD2 | ENSG00000134321 | Q8WXG1 | radical S-adenosyl methionine domain containing 2 | 298,69 | 2,04 | 0,44 | 4,68 | 2,89E-06 | 9,71E-04 |
| SERPING1 | ENSG00000149131 | P05155 | serpin family G member 1 | 106,77 | 1,98 | 0,42 | 4,75 | 2,01E-06 | 8,02E-04 |
| PTTG1 | ENSG00000164611 | O95997 | PTTG1 regulator of sister chromatid separation | 15,44 | 1,96 | 0,24 | 8,02 | 1,05E-15 | 9,37E-12 |
| ISG15 | ENSG00000187608 | P05161 | ISG15 ubiquitin like modifier | 198,86 | 1,94 | 0,42 | 4,59 | 4,47E-06 | 1,28E-03 |
| | ENSG00000224666 | | ETV7 and PTX1 antisense RNA 1 | 6,69 | 1,89 | 0,55 | 3,44 | 5,82E-04 | 2,24E-02 |
| SIGLEC1 | ENSG00000088827 | Q9BZZ2 | sialic acid binding Ig like lectin 1 | 20,73 | 1,73 | 0,42 | 4,16 | 3,15E-05 | 4,71E-03 |
| IGHG4 | ENSG00000211892 | P01861 | immunoglobulin heavy constant gamma 4 (G4m marker) | 16,91 | 1,71 | 0,39 | 4,41 | 1,04E-05 | 2,36E-03 |
| | ENSG00000210077 | | mitochondrially encoded tRNA-Val (GUN) | 428,73 | 1,70 | 0,39 | 4,39 | 1,11E-05 | 2,45E-03 |
| CMPK2 | ENSG00000134326 | Q5EBM0 | cytidine/uridine monophosphate kinase 2 | 100,85 | 1,66 | 0,42 | 3,96 | 7,59E-05 | 7,52E-03 |
| MZB1 | ENSG00000170476 | Q8WU39 | marginal zone B and B1 cell specific protein | 17,48 | 1,62 | 0,30 | 5,36 | 8,41E-08 | 7,35E-05 |
| BIRC5 | ENSG00000089685 | A0A0B4J1S3 | baculoviral IAP repeat containing 5 | 8,39 | 1,60 | 0,29 | 5,55 | 2,79E-08 | 3,31E-05 |
| FABP5 | ENSG00000164687 | Q01469 | fatty acid binding protein 5 | 9,41 | 1,53 | 0,27 | 5,78 | 7,40E-09 | 1,65E-05 |
| IFI44 | ENSG00000137965 | Q8TCB0 | interferon induced protein 44 | 134,89 | 1,52 | 0,39 | 3,92 | 8,97E-05 | 8,39E-03 |
| FAM111B | ENSG00000189057 | Q6SJ93 | FAM111 trypsin like peptidase B | 24,63 | 1,43 | 0,24 | 5,86 | 4,65E-09 | 1,38E-05 |
| TOP2A | ENSG00000131747 | P11388 | DNA topoisomerase II alpha | 79,11 | 1,43 | 0,17 | 8,20 | 2,42E-16 | 4,30E-12 |
| | ENSG00000211677 | | immunoglobulin lambda constant 2 | 276,85 | 1,43 | 0,26 | 5,57 | 2,59E-08 | 3,29E-05 |
| | ENSG00000211679 | | immunoglobulin lambda constant 3 (Kern-Oz+ marker) | 261,71 | 1,42 | 0,27 | 5,35 | 9,02E-08 | 7,35E-05 |
| IGHG1 | ENSG00000211896 | P01857 | immunoglobulin heavy constant gamma 1 (G1m marker) | 40,14 | 1,41 | 0,33 | 4,21 | 2,52E-05 | 4,15E-03 |
| | ENSG00000214653 | | heterogeneous nuclear ribonucleoprotein A3 pseudogene 3 | 10,21 | 1,40 | 0,44 | 3,15 | 1,61E-03 | 3,91E-02 |
| LY6E | ENSG00000160932 | E5RGI6 | lymphocyte antigen 6 family member E | 83,32 | 1,40 | 0,29 | 4,82 | 1,42E-06 | 6,67E-04 |
| | ENSG00000209082 | | mitochondrially encoded tRNA-Leu (UUA/G) 1 | 224,75 | 1,38 | 0,32 | 4,24 | 2,28E-05 | 3,87E-03 |
| FBXO39 | ENSG00000177294 | Q8N4B4 | F-box protein 39 | 12,17 | 1,37 | 0,35 | 3,96 | 7,35E-05 | 7,50E-03 |
| HERC5 | ENSG00000138646 | Q9UII4 | HECT and RLD domain containing E3 ubiquitin protein ligase 5 | 365,89 | 1,37 | 0,36 | 3,75 | 1,75E-04 | 1,26E-02 |
| EPSTI1 | ENSG00000133106 | Q96J88 | epithelial stromal interaction 1 | 864,53 | 1,37 | 0,34 | 4,07 | 4,64E-05 | 5,74E-03 |
| | ENSG00000239039 | | small nucleolar RNA | 244,92 | 1,36 | 0,27 | 4,99 | 5,95E-07 | 3,65E-04 |
| MT1E | ENSG00000169715 | P04732 | metallothionein 1E | 22,95 | 1,34 | 0,33 | 4,12 | 3,82E-05 | 5,28E-03 |
| CDC20 | ENSG00000117399 | Q12834 | cell division cycle 20 | 5,34 | 1,29 | 0,29 | 4,36 | 1,28E-05 | 2,58E-03 |
| GBP1 | ENSG00000117228 | P32455 | guanylate binding protein 1 | 701,90 | 1,28 | 0,32 | 4,01 | 6,03E-05 | 6,76E-03 |
| RPL9 | ENSG00000163682 | Q53Z07 | ribosomal protein L9 | 426,17 | 1,28 | 0,26 | 4,84 | 1,31E-06 | 6,48E-04 |
| IGHA2 | ENSG00000211890 | P01877 | immunoglobulin heavy constant alpha 2 (A2m marker) | 26,69 | 1,24 | 0,27 | 4,55 | 5,38E-06 | 1,47E-03 |
| IGHA1 | ENSG00000211895 | P01876 | immunoglobulin heavy constant alpha 1 | 97,89 | 1,23 | 0,26 | 4,75 | 2,07E-06 | 8,02E-04 |
| H4C1 | ENSG00000197238 | P62805 | H4 clustered histone 11 | 7,34 | 1,22 | 0,26 | 4,74 | 2,12E-06 | 8,02E-04 |
| MX1 | ENSG00000157601 | P20591 | MX dynamin like GTPase 1 | 453,81 | 1,21 | 0,36 | 3,39 | 7,08E-04 | 2,45E-02 |
| GBP5 | ENSG00000154451 | Q96PP8 | guanylate binding protein 5 | 81,35 | 1,21 | 0,34 | 3,53 | 4,20E-04 | 1,91E-02 |
| RTP4 | ENSG00000136514 | Q96DX8 | receptor transporter protein 4 | 31,16 | 1,21 | 0,31 | 3,85 | 1,18E-04 | 9,93E-03 |
| ETV7 | ENSG00000010030 | Q9Y603 | ETS variant transcription factor 7 | 36,79 | 1,20 | 0,38 | 3,20 | 1,38E-03 | 3,57E-02 |
| | ENSG00000211897 | | immunoglobulin heavy constant gamma 3 (G3m marker) | 36,97 | 1,20 | 0,31 | 3,87 | 1,09E-04 | 9,44E-03 |
| STMN1 | ENSG00000117632 | P16949 | stathmin 1 | 88,06 | 1,19 | 0,17 | 6,94 | 4,00E-12 | 2,37E-08 |
| MKI67 | ENSG00000148773 | P46013 | marker of proliferation Ki-67 | 84,79 | 1,18 | 0,20 | 6,04 | 1,55E-09 | 5,52E-06 |
| | ENSG00000225492 | | guanylate binding protein 1 pseudogene 1 | 157,86 | 1,17 | 0,34 | 3,44 | 5,92E-04 | 2,26E-02 |
| SMTNL1 | ENSG00000214872 | A8MU46 | smoothelin like 1 | 89,92 | 1,13 | 0,25 | 4,56 | 5,10E-06 | 1,44E-03 |
| C1QA | ENSG00000173372 | P02745 | complement C1q A chain | 18,20 | 1,12 | 0,36 | 3,11 | 1,85E-03 | 4,31E-02 |
| H4C1 | ENSG00000273542 | P62805 | H4 clustered histone 12 | 5,59 | 1,12 | 0,31 | 3,60 | 3,17E-04 | 1,69E-02 |
| AURKB | ENSG00000178999 | Q96GD4 | aurora kinase B | 10,45 | 1,11 | 0,26 | 4,35 | 1,33E-05 | 2,64E-03 |

| | | | | | | | | | |
|---|---|---|---|---|---|---|---|---|---|
| | ENSG00000254258 | | novel transcript | 7,77 | 1,10 | 0,30 | 3,66 | 2,49E-04 | 1,44E-02 |
| XAF1 | ENSG00000132530 | Q6GPH4 | XIAP associated factor 1 | 83,59 | 1,10 | 0,26 | 4,27 | 1,96E-05 | 3,60E-03 |
| CDCA7 | ENSG00000144354 | Q9BWT1 | cell division cycle associated 7 | 7,20 | 1,09 | 0,26 | 4,11 | 4,02E-05 | 5,33E-03 |
| IFIT5 | ENSG00000152778 | Q13325 | interferon induced protein with tetratricopeptide repeats 5 | 53,69 | 1,07 | 0,26 | 4,11 | 3,89E-05 | 5,32E-03 |
| | ENSG00000210184 | | mitochondrially encoded tRNA-Ser (AGU/C) 2 | 24,29 | 1,07 | 0,34 | 3,18 | 1,47E-03 | 3,70E-02 |
| TTK | ENSG00000112742 | P33981 | TTK protein kinase | 5,49 | 1,07 | 0,24 | 4,43 | 9,60E-06 | 2,22E-03 |
| | ENSG00000240087 | | ribosomal protein SA pseudogene 12 | 7,19 | 1,06 | 0,34 | 3,14 | 1,71E-03 | 4,07E-02 |
| USP18 | ENSG00000184979 | Q9UMW8 | ubiquitin specific peptidase 18 | 47,37 | 1,04 | 0,30 | 3,50 | 4,70E-04 | 2,01E-02 |
| GGH | ENSG00000137563 | Q92820 | gamma-glutamyl hydrolase | 6,40 | 1,03 | 0,25 | 4,14 | 3,45E-05 | 4,96E-03 |
| ASPM | ENSG00000066279 | Q8IZT6 | assembly factor for spindle microtubules | 34,50 | 1,01 | 0,19 | 5,34 | 9,08E-08 | 7,35E-05 |
| MCM10 | ENSG00000065328 | Q7L590 | minichromosome maintenance 10 replication initiation factor | 14,79 | 1,01 | 0,18 | 5,60 | 2,10E-08 | 3,03E-05 |
| KIFC1 | ENSG00000237649 | Q9BW19 | kinesin family member C1 | 15,06 | 1,01 | 0,18 | 5,51 | 3,49E-08 | 3,86E-05 |
| CKS1B | ENSG00000173207 | P61024 | CDC28 protein kinase regulatory subunit 1B | 20,83 | 1,00 | 0,18 | 5,68 | 1,39E-08 | 2,47E-05 |
| HMGB3 | ENSG00000029993 | O15347 | high mobility group box 3 | 5,79 | 1,00 | 0,24 | 4,21 | 2,59E-05 | 4,22E-03 |
| GZMA | ENSG00000145649 | P12544 | granzyme A | 159,90 | 1,00 | 0,21 | 4,71 | 2,50E-06 | 8,74E-04 |
| RPS15A | ENSG00000134419 | P62244 | ribosomal protein S15a | 431,62 | 0,96 | 0,23 | 4,23 | 2,31E-05 | 3,87E-03 |
| SIT1 | ENSG00000137078 | Q9Y3P8 | signaling threshold regulating transmembrane adaptor 1 | 57,62 | 0,96 | 0,19 | 5,03 | 4,78E-07 | 3,04E-04 |
| | ENSG00000214273 | | angiogenic factor with G-patch and FHA domains 1 pseudogene 1 | 14,25 | 0,96 | 0,27 | 3,56 | 3,65E-04 | 1,80E-02 |
| CENPF | ENSG00000117724 | P49454 | centromere protein F | 66,09 | 0,95 | 0,16 | 5,80 | 6,73E-09 | 1,65E-05 |
| LAP3 | ENSG00000002549 | P28838 | leucine aminopeptidase 3 | 88,70 | 0,93 | 0,26 | 3,59 | 3,26E-04 | 1,70E-02 |
| | ENSG00000233435 | | angiogenic factor with G-patch and FHA domains 1 pseudogene 2 | 16,47 | 0,93 | 0,27 | 3,38 | 7,23E-04 | 2,47E-02 |
| SAMD9L | ENSG00000177409 | Q8IVG5 | sterile alpha motif domain containing 9 like | 874,80 | 0,92 | 0,26 | 3,59 | 3,28E-04 | 1,70E-02 |
| OASL | ENSG00000135114 | Q15646 | 2'-5'-oligoadenylate synthetase like | 109,24 | 0,92 | 0,27 | 3,35 | 8,20E-04 | 2,62E-02 |
| C4orf48 | ENSG00000243449 | Q5BLP8 | chromosome 4 open reading frame 48 | 15,87 | 0,92 | 0,22 | 4,23 | 2,30E-05 | 3,87E-03 |
| | ENSG00000210107 | | mitochondrially encoded tRNA-Gln (CAA/G) | 278,13 | 0,92 | 0,25 | 3,71 | 2,11E-04 | 1,35E-02 |
| HMMR | ENSG00000072571 | O75330 | hyaluronan mediated motility receptor | 8,60 | 0,91 | 0,23 | 3,98 | 6,77E-05 | 7,25E-03 |
| | ENSG00000267696 | | endogenous retrovirus group K member 28 | 43,71 | 0,90 | 0,23 | 3,87 | 1,07E-04 | 9,41E-03 |
| PSME2 | ENSG00000100911 | Q86SZ7 | proteasome activator subunit 2 | 175,62 | 0,90 | 0,23 | 3,89 | 1,02E-04 | 9,13E-03 |
| MCM4 | ENSG00000104738 | P33991 | minichromosome maintenance complex component 4 | 18,58 | 0,90 | 0,17 | 5,17 | 2,39E-07 | 1,70E-04 |
| CCNB1 | ENSG00000134057 | P14635 | cyclin B1 | 7,03 | 0,90 | 0,21 | 4,25 | 2,15E-05 | 3,76E-03 |
| VAMP5 | ENSG00000168899 | O95183 | vesicle associated membrane protein 5 | 39,08 | 0,89 | 0,20 | 4,41 | 1,05E-05 | 2,36E-03 |
| PERP | ENSG00000112378 | Q96FX8 | p53 apoptosis effector related to PMP22 | 8,10 | 0,88 | 0,24 | 3,66 | 2,56E-04 | 1,47E-02 |
| RMI2 | ENSG00000175643 | Q96E14 | RecQ mediated genome instability 2 | 69,71 | 0,87 | 0,26 | 3,39 | 7,00E-04 | 2,45E-02 |
| | ENSG00000231873 | | novel transcript | 38,11 | 0,87 | 0,26 | 3,40 | 6,83E-04 | 2,40E-02 |
| CETP | ENSG00000087237 | P11597 | cholesteryl ester transfer protein | 239,62 | 0,87 | 0,20 | 4,33 | 1,47E-05 | 2,85E-03 |
| CDC6 | ENSG00000094804 | Q99741 | cell division cycle 6 | 6,02 | 0,86 | 0,24 | 3,51 | 4,55E-04 | 1,97E-02 |
| | ENSG00000210049 | | mitochondrially encoded tRNA-Phe (UUU/C) | 172,44 | 0,85 | 0,25 | 3,36 | 7,79E-04 | 2,58E-02 |
| | ENSG00000244558 | | KCNK15 and WISP2 antisense RNA 1 | 7,77 | 0,85 | 0,22 | 3,83 | 1,28E-04 | 1,04E-02 |
| CD52 | ENSG00000169442 | P31358 | CD52 molecule | 773,38 | 0,85 | 0,19 | 4,50 | 6,91E-06 | 1,79E-03 |
| NAP1L2 | ENSG00000186462 | Q9ULW6 | nucleosome assembly protein 1 like 2 | 12,14 | 0,85 | 0,28 | 3,06 | 2,18E-03 | 4,76E-02 |
| RRM2 | ENSG00000171848 | P31350 | ribonucleotide reductase regulatory subunit M2 | 55,17 | 0,84 | 0,15 | 5,74 | 9,35E-09 | 1,85E-05 |
| LAG3 | ENSG00000089692 | P18627 | lymphocyte activating 3 | 13,09 | 0,84 | 0,25 | 3,33 | 8,78E-04 | 2,71E-02 |
| E2F7 | ENSG00000165891 | Q96AV8 | E2F transcription factor 7 | 9,50 | 0,84 | 0,21 | 4,06 | 4,89E-05 | 5,80E-03 |
| ISOC2 | ENSG00000063241 | Q96AB3 | isochorismatase domain containing 2 | 9,39 | 0,84 | 0,21 | 4,03 | 5,65E-05 | 6,41E-03 |
| CTSL | ENSG00000135047 | P07711 | cathepsin L | 13,44 | 0,84 | 0,23 | 3,70 | 2,16E-04 | 1,36E-02 |
| ACP2 | ENSG00000134575 | B7Z7D2 | acid phosphatase 2 | 103,79 | 0,83 | 0,25 | 3,40 | 6,77E-04 | 2,39E-02 |
| | ENSG00000229119 | | 60S acidic ribosomal protein (RPLP0) pseudogene | 78,00 | 0,81 | 0,17 | 4,81 | 1,48E-06 | 6,75E-04 |
| | ENSG00000285080 | | novel transcript | 44,89 | 0,81 | 0,25 | 3,29 | 1,00E-03 | 2,93E-02 |
| RPL39 | ENSG00000198918 | P62891 | ribosomal protein L39 | 860,33 | 0,81 | 0,19 | 4,20 | 2,62E-05 | 4,24E-03 |
| PFDN6 | ENSG00000204220 | Q5STK2 | prefoldin subunit 6 [Source:HGNC Symbol;Acc:HGNC:4926] | 35,53 | 0,80 | 0,16 | 4,99 | 6,14E-07 | 3,65E-04 |

| | | | | | | | | | |
|---|---|---|---|---|---|---|---|---|---|
| | ENSG00000230721 | | ribosomal protein L17 (RPL17) pseudogene | 9,38 | 0,80 | 0,25 | 3,23 | 1,22E-03 | 3,34E-02 |
| PLAAT4 | ENSG00000133321 | Q9UL19 | phospholipase A and acyltransferase 4 | 337,50 | 0,80 | 0,14 | 5,64 | 1,73E-08 | 2,80E-05 |
| OLFML3 | ENSG00000116774 | B4DNG0 | olfactomedin like 3 | 6,75 | 0,80 | 0,26 | 3,09 | 2,01E-03 | 4,54E-02 |
| | ENSG00000226715 | | long intergenic non-protein coding RNA 1709 | 73,49 | 0,80 | 0,22 | 3,66 | 2,48E-04 | 1,44E-02 |
| H1-2 | ENSG00000187837 | P16403 | H1.2 linker histone | 552,88 | 0,79 | 0,18 | 4,36 | 1,29E-05 | 2,58E-03 |
| | ENSG00000259642 | | ST20 antisense RNA 1 | 6,39 | 0,79 | 0,23 | 3,36 | 7,83E-04 | 2,59E-02 |
| TIMM10 | ENSG00000134809 | P62072 | translocase of inner mitochondrial membrane 10 | 30,92 | 0,78 | 0,22 | 3,64 | 2,74E-04 | 1,52E-02 |
| | ENSG00000231663 | | COA6 antisense RNA 1 | 6,45 | 0,78 | 0,23 | 3,42 | 6,36E-04 | 2,34E-02 |
| BATF3 | ENSG00000123685 | Q9NR55 | basic leucine zipper ATF-like transcription factor 3 | 8,28 | 0,78 | 0,26 | 3,05 | 2,31E-03 | 4,93E-02 |
| CISD3 | ENSG00000277972 | P0C7P0 | CDGSH iron sulfur domain 3 | 30,03 | 0,77 | 0,14 | 5,51 | 3,69E-08 | 3,86E-05 |
| UQCRQ | ENSG00000164405 | O14949 | ubiquinol-cytochrome c reductase complex III subunit VII | 189,16 | 0,77 | 0,16 | 4,82 | 1,42E-06 | 6,67E-04 |
| SDF2L1 | ENSG00000128228 | Q9HCN8 | stromal cell derived factor 2 like 1 | 87,53 | 0,77 | 0,18 | 4,36 | 1,29E-05 | 2,58E-03 |
| | ENSG00000235576 | | long intergenic non-protein coding RNA 1871 | 195,94 | 0,77 | 0,24 | 3,20 | 1,36E-03 | 3,57E-02 |
| RPL27 | ENSG00000131469 | A0A024R1V4 | ribosomal protein L27 | 416,72 | 0,77 | 0,21 | 3,60 | 3,15E-04 | 1,68E-02 |
| IFI35 | ENSG00000068079 | P80217 | interferon induced protein 35 | 59,99 | 0,77 | 0,23 | 3,30 | 9,60E-04 | 2,86E-02 |
| GBP4 | ENSG00000162654 | Q96PP9 | guanylate binding protein 4 | 306,50 | 0,77 | 0,24 | 3,16 | 1,58E-03 | 3,89E-02 |
| | ENSG00000256582 | | long intergenic non-protein coding RNA 2390 | 5,51 | 0,77 | 0,24 | 3,14 | 1,67E-03 | 4,01E-02 |
| TCF19 | ENSG00000137310 | Q9Y242 | transcription factor 19 | 9,48 | 0,76 | 0,22 | 3,49 | 4,82E-04 | 2,02E-02 |
| MAGEE1 | ENSG00000198934 | Q9HCI5 | MAGE family member E1 | 5,90 | 0,76 | 0,23 | 3,29 | 9,90E-04 | 2,92E-02 |
| DBI | ENSG00000155368 | P07108 | diazepam binding inhibitor | 186,53 | 0,76 | 0,18 | 4,17 | 3,05E-05 | 4,71E-03 |
| HINT1 | ENSG00000169567 | D6RC06 | histidine triad nucleotide binding protein 1 | 612,82 | 0,75 | 0,20 | 3,66 | 2,49E-04 | 1,44E-02 |
| BOLA2 | ENSG00000169627 | Q9H3K6 | bolA family member 2B | 87,08 | 0,74 | 0,16 | 4,77 | 1,80E-06 | 7,63E-04 |
| DUSP5 | ENSG00000138166 | Q16690 | dual specificity phosphatase 5 | 29,82 | 0,74 | 0,18 | 4,25 | 2,16E-05 | 3,76E-03 |
| CEP55 | ENSG00000138180 | Q53EZ4 | centrosomal protein 55 | 16,60 | 0,74 | 0,18 | 4,12 | 3,79E-05 | 5,28E-03 |
| DDX60 | ENSG00000137628 | Q8IY21 | DExD/H-box helicase 60 | 344,64 | 0,74 | 0,23 | 3,22 | 1,29E-03 | 3,46E-02 |
| APOL6 | ENSG00000221963 | Q9BWW8 | apolipoprotein L6 | 628,89 | 0,74 | 0,21 | 3,47 | 5,21E-04 | 2,12E-02 |
| DTL | ENSG00000143476 | Q9NZJ0 | denticleless E3 ubiquitin protein ligase homolog | 29,27 | 0,74 | 0,15 | 4,94 | 7,92E-07 | 4,27E-04 |
| GINS2 | ENSG00000131153 | Q9Y248 | GINS complex subunit 2 | 8,87 | 0,73 | 0,21 | 3,50 | 4,60E-04 | 1,97E-02 |
| ANTKMT | ENSG00000103254 | Q9BQD7 | adenine nucleotide translocase lysine methyltransferase | 6,57 | 0,73 | 0,23 | 3,20 | 1,37E-03 | 3,57E-02 |
| FBXO5 | ENSG00000112029 | Q9UKT4 | F-box protein 5 | 12,20 | 0,73 | 0,19 | 3,84 | 1,21E-04 | 1,00E-02 |
| H4C1 | ENSG00000197061 | P62805 | H4 clustered histone 3 | 941,47 | 0,73 | 0,21 | 3,43 | 6,14E-04 | 2,30E-02 |
| TXNDC17 | ENSG00000129235 | Q9BRA2 | thioredoxin domain containing 17 | 42,48 | 0,72 | 0,17 | 4,16 | 3,12E-05 | 4,71E-03 |
| | ENSG00000255571 | | MIR9-3 host gene | 6,32 | 0,72 | 0,22 | 3,30 | 9,69E-04 | 2,87E-02 |
| RPA3 | ENSG00000106399 | P35244 | replication protein A3 | 35,48 | 0,72 | 0,20 | 3,54 | 4,07E-04 | 1,88E-02 |
| | ENSG00000243819 | | RNA | 10,39 | 0,72 | 0,19 | 3,86 | 1,11E-04 | 9,56E-03 |
| ROMO1 | ENSG00000125995 | P60602 | reactive oxygen species modulator 1 | 148,18 | 0,72 | 0,22 | 3,24 | 1,17E-03 | 3,28E-02 |
| ZBP1 | ENSG00000124256 | Q9H171 | Z-DNA binding protein 1 | 237,72 | 0,72 | 0,22 | 3,21 | 1,31E-03 | 3,48E-02 |
| UBE2L6 | ENSG00000156587 | O14933 | ubiquitin conjugating enzyme E2 L6 | 255,02 | 0,72 | 0,22 | 3,30 | 9,62E-04 | 2,86E-02 |
| NXT2 | ENSG00000101888 | Q9NPJ8 | nuclear transport factor 2 like export factor 2 | 7,77 | 0,71 | 0,19 | 3,67 | 2,43E-04 | 1,44E-02 |
| RPS7 | ENSG00000171863 | P62081 | ribosomal protein S7 | 364,25 | 0,71 | 0,20 | 3,65 | 2,66E-04 | 1,50E-02 |
| EEF1B2 | ENSG00000114942 | P24534 | eukaryotic translation elongation factor 1 beta 2 | 228,95 | 0,71 | 0,21 | 3,39 | 7,10E-04 | 2,45E-02 |
| H1-4 | ENSG00000168298 | P10412 | H1.4 linker histone | 85,55 | 0,71 | 0,23 | 3,08 | 2,07E-03 | 4,62E-02 |
| CDCA2 | ENSG00000184661 | Q69YH5 | cell division cycle associated 2 | 13,52 | 0,71 | 0,20 | 3,58 | 3,38E-04 | 1,72E-02 |
| EXO1 | ENSG00000174371 | Q9UQ84 | exonuclease 1 | 9,80 | 0,70 | 0,21 | 3,32 | 9,12E-04 | 2,77E-02 |
| COX7B | ENSG00000131174 | P24311 | cytochrome c oxidase subunit 7B | 41,78 | 0,70 | 0,20 | 3,49 | 4,77E-04 | 2,02E-02 |
| RPLP0 | ENSG00000089157 | P05388 | ribosomal protein lateral stalk subunit P0 | 683,50 | 0,70 | 0,15 | 4,65 | 3,24E-06 | 1,06E-03 |
| CLSPN | ENSG00000092853 | Q9HAW4 | claspin | 44,19 | 0,70 | 0,15 | 4,71 | 2,50E-06 | 8,74E-04 |
| RPSA | ENSG00000168028 | P08865 | ribosomal protein SA | 489,37 | 0,70 | 0,17 | 4,06 | 4,86E-05 | 5,80E-03 |
| MAGEH1 | ENSG00000187601 | Q9H213 | MAGE family member H1 | 19,51 | 0,70 | 0,19 | 3,76 | 1,70E-04 | 1,25E-02 |
| NCAPH | ENSG00000121152 | Q15003 | non-SMC condensin I complex subunit H | 23,35 | 0,70 | 0,16 | 4,49 | 7,23E-06 | 1,84E-03 |

| | | | | | | | | | |
|---|---|---|---|---|---|---|---|---|---|
| ZNF688 | ENSG00000229809 | P0C7X2 | zinc finger protein 688 | 23,19 | 0,69 | 0,15 | 4,59 | 4,42E-06 | 1,28E-03 |
| ESCO2 | ENSG00000171320 | Q56NI9 | establishment of sister chromatid cohesion N-acetyltransferase 2 | 11,92 | 0,69 | 0,20 | 3,48 | 4,92E-04 | 2,04E-02 |
| PSMG1 | ENSG00000183527 | O95456 | proteasome assembly chaperone 1 | 9,21 | 0,69 | 0,20 | 3,45 | 5,69E-04 | 2,21E-02 |
| BMI1 | ENSG00000168283 | P35226 | BMI1 proto-oncogene | 14,92 | 0,68 | 0,20 | 3,42 | 6,24E-04 | 2,32E-02 |
| MT1F | ENSG00000198417 | P04733 | metallothionein 1F | 30,17 | 0,68 | 0,16 | 4,15 | 3,28E-05 | 4,83E-03 |
| CXCR3 | ENSG00000186810 | P49682 | C-X-C motif chemokine receptor 3 | 27,67 | 0,68 | 0,19 | 3,50 | 4,58E-04 | 1,97E-02 |
| TIMM8B | ENSG00000150779 | Q9Y5J9 | translocase of inner mitochondrial membrane 8 homolog B | 19,74 | 0,68 | 0,19 | 3,56 | 3,66E-04 | 1,80E-02 |
| CCNQ | ENSG00000262919 | Q8N1B3 | cyclin Q | 37,85 | 0,67 | 0,20 | 3,43 | 6,11E-04 | 2,30E-02 |
| | ENSG00000287697 | | novel transcript | 8,30 | 0,67 | 0,19 | 3,52 | 4,27E-04 | 1,91E-02 |
| CENPE | ENSG00000138778 | Q02224 | centromere protein E | 56,12 | 0,67 | 0,12 | 5,47 | 4,51E-08 | 4,46E-05 |
| CDKN2C | ENSG00000123080 | P42773 | cyclin dependent kinase inhibitor 2C | 8,02 | 0,67 | 0,22 | 3,06 | 2,21E-03 | 4,79E-02 |
| | ENSG00000286848 | | novel transcript | 19,68 | 0,67 | 0,21 | 3,27 | 1,09E-03 | 3,12E-02 |
| SAMD9 | ENSG00000205413 | Q5K651 | sterile alpha motif domain containing 9 | 425,82 | 0,67 | 0,19 | 3,44 | 5,77E-04 | 2,23E-02 |
| PTRHD1 | ENSG00000184924 | Q6GMV3 | peptidyl-tRNA hydrolase domain containing 1 | 25,42 | 0,67 | 0,19 | 3,49 | 4,83E-04 | 2,02E-02 |
| CHST10 | ENSG00000115526 | O43529 | carbohydrate sulfotransferase 10 | 9,81 | 0,66 | 0,20 | 3,36 | 7,89E-04 | 2,59E-02 |
| TRAPPC2L | ENSG00000167515 | Q9UL33 | trafficking protein particle complex 2 like | 34,82 | 0,66 | 0,16 | 4,09 | 4,29E-05 | 5,51E-03 |
| CD8A | ENSG00000153563 | P01732 | CD8a molecule | 129,00 | 0,66 | 0,19 | 3,44 | 5,86E-04 | 2,25E-02 |
| MRPL17 | ENSG00000158042 | Q9NRX2 | mitochondrial ribosomal protein L17 | 15,99 | 0,66 | 0,19 | 3,42 | 6,33E-04 | 2,33E-02 |
| MRPL23 | ENSG00000214026 | Q16540 | mitochondrial ribosomal protein L23 | 25,69 | 0,66 | 0,18 | 3,67 | 2,46E-04 | 1,44E-02 |
| RPS6KB2 | ENSG00000175634 | Q9UBS0 | ribosomal protein S6 kinase B2 | 13,86 | 0,66 | 0,18 | 3,56 | 3,71E-04 | 1,81E-02 |
| RPS29 | ENSG00000213741 | P62273 | ribosomal protein S29 | 8998,61 | 0,66 | 0,19 | 3,46 | 5,42E-04 | 2,15E-02 |
| RPS18 | ENSG00000231500 | P62269 | ribosomal protein S18 | 1401,14 | 0,65 | 0,20 | 3,25 | 1,14E-03 | 3,23E-02 |
| RDH14 | ENSG00000240857 | Q9HBH5 | retinol dehydrogenase 14 | 17,36 | 0,65 | 0,19 | 3,42 | 6,22E-04 | 2,32E-02 |
| ZNF408 | ENSG00000175213 | Q9H9D4 | zinc finger protein 408 | 10,35 | 0,65 | 0,17 | 3,84 | 1,24E-04 | 1,02E-02 |
| TIFA | ENSG00000145365 | Q96CG3 | TRAF interacting protein with forkhead associated domain | 45,42 | 0,65 | 0,16 | 4,03 | 5,52E-05 | 6,29E-03 |
| STAT2 | ENSG00000170581 | R9QE65 | signal transducer and activator of transcription 2 | 196,69 | 0,65 | 0,19 | 3,35 | 7,97E-04 | 2,60E-02 |
| DNPH1 | ENSG00000112667 | O43598 | 2'-deoxynucleoside 5'-phosphate N-hydrolase 1 | 47,59 | 0,65 | 0,12 | 5,31 | 1,08E-07 | 8,40E-05 |
| TRIM68 | ENSG00000167333 | Q6AZZ1 | tripartite motif containing 68 | 7,96 | 0,65 | 0,18 | 3,52 | 4,37E-04 | 1,93E-02 |
| BUB1 | ENSG00000169679 | O43683 | BUB1 mitotic checkpoint serine/threonine kinase | 16,50 | 0,64 | 0,16 | 3,93 | 8,46E-05 | 8,18E-03 |
| PDCD5 | ENSG00000105185 | O14737 | programmed cell death 5 | 46,01 | 0,64 | 0,19 | 3,36 | 7,74E-04 | 2,58E-02 |
| RPL41 | ENSG00000229117 | P62945 | ribosomal protein L41 | 3867,40 | 0,64 | 0,17 | 3,70 | 2,18E-04 | 1,36E-02 |
| RAN | ENSG00000132341 | B4DV51 | µ | 347,28 | 0,64 | 0,19 | 3,35 | 8,07E-04 | 2,61E-02 |
| SLAMF7 | ENSG00000026751 | Q9NQ25 | SLAM family member 7 | 198,35 | 0,64 | 0,15 | 4,39 | 1,14E-05 | 2,47E-03 |
| | ENSG00000236552 | | ribosomal protein L13a pseudogene 5 | 481,45 | 0,64 | 0,15 | 4,13 | 3,57E-05 | 5,04E-03 |
| NDUFA4 | ENSG00000189043 | O00483 | NDUFA4 mitochondrial complex associated | 95,05 | 0,64 | 0,20 | 3,13 | 1,73E-03 | 4,10E-02 |
| | ENSG00000239636 | | TFAP2E antisense RNA 1 | 14,99 | 0,63 | 0,16 | 4,00 | 6,39E-05 | 6,97E-03 |
| | ENSG00000273061 | | CDC37L1 divergent transcript | 12,97 | 0,63 | 0,19 | 3,39 | 7,05E-04 | 2,45E-02 |
| NCBP2AS2 | ENSG00000270170 | Q69YL0 | NCBP2 antisense 2 (head to head) | 39,92 | 0,62 | 0,15 | 4,14 | 3,48E-05 | 4,96E-03 |
| IFI27L2 | ENSG00000119632 | Q9H2X8 | interferon alpha inducible protein 27 like 2 | 46,12 | 0,62 | 0,17 | 3,58 | 3,43E-04 | 1,74E-02 |
| PDRG1 | ENSG00000088356 | Q9NUG6 | p53 and DNA damage regulated 1 | 28,14 | 0,62 | 0,15 | 4,17 | 3,05E-05 | 4,71E-03 |
| | ENSG00000234498 | | ribosomal protein L13a pseudogene 20 | 335,92 | 0,62 | 0,19 | 3,23 | 1,24E-03 | 3,38E-02 |
| | ENSG00000232956 | | small nucleolar RNA host gene 15 | 12,26 | 0,62 | 0,18 | 3,46 | 5,31E-04 | 2,13E-02 |
| RPL35 | ENSG00000136942 | P42766 | ribosomal protein L35 | 201,40 | 0,62 | 0,18 | 3,48 | 5,04E-04 | 2,07E-02 |
| RPL27A | ENSG00000166441 | P46776 | ribosomal protein L27a | 2490,46 | 0,61 | 0,14 | 4,40 | 1,06E-05 | 2,37E-03 |
| TRIM69 | ENSG00000185880 | Q86WT6 | tripartite motif containing 69 | 138,82 | 0,61 | 0,13 | 4,80 | 1,60E-06 | 7,11E-04 |
| AKIP1 | ENSG00000166452 | Q9NQ31 | A-kinase interacting protein 1 | 13,34 | 0,61 | 0,18 | 3,38 | 7,15E-04 | 2,46E-02 |
| COQ10A | ENSG00000135469 | Q96MF6 | coenzyme Q10A | 8,17 | 0,61 | 0,20 | 3,07 | 2,13E-03 | 4,70E-02 |
| SCOC | ENSG00000153130 | A0A0C4DGB0 | short coiled-coil protein | 20,09 | 0,61 | 0,20 | 3,11 | 1,86E-03 | 4,31E-02 |
| TMEM258 | ENSG00000134825 | P61165 | transmembrane protein 258 | 198,64 | 0,61 | 0,16 | 3,84 | 1,24E-04 | 1,02E-02 |
| TMEM147 | ENSG00000105677 | Q9BVK8 | transmembrane protein 147 | 62,18 | 0,61 | 0,14 | 4,27 | 1,99E-05 | 3,61E-03 |

### *Supplementary Table S3: Top 200 most upregulated DEGs during HZ compared to controls*

| uniprot_gn_symbol | ensembl_gene_id | uniprot_gn_id | description | baseMean | log2 FoldChange | lfcSE | stat | p-value | Adj p-value |
|---|---|---|---|---|---|---|---|---|---|
| IFI27 | ENSG00000165949 | P40305 | interferon alpha inducible protein 27 | 75,50 | 2,62 | 0,48 | 5,45 | 5,16E-08 | 5,82E-05 |
| | ENSG00000253755 | | immunoglobulin heavy constant gamma P (non-functional) | 12,39 | 2,36 | 0,41 | 5,76 | 8,62E-09 | 1,63E-05 |
| ZWINT | ENSG00000122952 | O95229 | ZW10 interacting kinetochore protein | 4,85 | 2,08 | 0,37 | 5,69 | 1,24E-08 | 1,80E-05 |
| C4BPA | ENSG00000123838 | P04003 | complement component 4 binding protein | 22,85 | 2,01 | 0,44 | 4,55 | 5,34E-06 | 2,05E-03 |
| CDK1 | ENSG00000170312 | P06493 | cyclin dependent kinase 1 | 3,95 | 1,81 | 0,36 | 5,03 | 4,88E-07 | 3,34E-04 |
| FAM111B | ENSG00000189057 | Q6SJ93 | FAM111 trypsin like peptidase B | 24,63 | 1,63 | 0,24 | 6,78 | 1,17E-11 | 5,61E-08 |
| IFI44L | ENSG00000137959 | Q53G44 | interferon induced protein 44 like | 273,91 | 1,60 | 0,46 | 3,48 | 5,03E-04 | 3,08E-02 |
| IGHG4 | ENSG00000211892 | P01861 | immunoglobulin heavy constant gamma 4 (G4m marker) | 16,91 | 1,60 | 0,38 | 4,23 | 2,38E-05 | 5,86E-03 |
| BIRC5 | ENSG00000089685 | A0A0B4J1S3 | baculoviral IAP repeat containing 5 | 8,39 | 1,54 | 0,28 | 5,49 | 3,92E-08 | 5,02E-05 |
| | ENSG00000210077 | | mitochondrially encoded tRNA-Val (GUN) | 428,73 | 1,50 | 0,38 | 3,95 | 7,66E-05 | 1,07E-02 |
| H2AC14 | ENSG00000276368 | Q99878 | H2A clustered histone 14 | 5,08 | 1,50 | 0,34 | 4,34 | 1,41E-05 | 4,31E-03 |
| HJURP | ENSG00000123485 | Q8NCD3 | Holliday junction recognition protein | 3,70 | 1,45 | 0,30 | 4,85 | 1,22E-06 | 6,70E-04 |
| FABP5 | ENSG00000164687 | Q01469 | fatty acid binding protein 5 | 9,41 | 1,40 | 0,26 | 5,45 | 5,06E-08 | 5,82E-05 |
| IGHV1OR15-1 | ENSG00000270505 | A0A075B7D0 | immunoglobulin heavy variable 1/OR15-1 (non-functional) | 4,23 | 1,38 | 0,33 | 4,23 | 2,37E-05 | 5,86E-03 |
| RPL9 | ENSG00000163682 | Q53Z07 | ribosomal protein L9 | 426,17 | 1,34 | 0,26 | 5,20 | 2,04E-07 | 1,70E-04 |
| PTTG1 | ENSG00000164611 | O95997 | PTTG1 regulator of sister chromatid separation | 15,44 | 1,34 | 0,23 | 5,82 | 5,78E-09 | 1,23E-05 |
| TOP2A | ENSG00000131747 | P11388 | DNA topoisomerase II alpha | 79,11 | 1,33 | 0,17 | 7,85 | 4,13E-15 | 7,93E-11 |
| IGHV1OR15-1 | ENSG00000281179 | A0A075B7D0 | novel gene identicle to IGHV1OR15-1 | 4,28 | 1,33 | 0,32 | 4,15 | 3,27E-05 | 7,05E-03 |
| PDILT | ENSG00000169340 | Q8N807 | protein disulfide isomerase like | 4,05 | 1,25 | 0,31 | 4,09 | 4,29E-05 | 8,23E-03 |
| S100B | ENSG00000160307 | A8MRB1 | S100 calcium binding protein B | 11,49 | 1,23 | 0,34 | 3,66 | 2,48E-04 | 2,11E-02 |
| CALHM6 | ENSG00000188820 | Q5R3K3 | calcium homeostasis modulator family member | 5,10 | 1,22 | 0,37 | 3,35 | 8,11E-04 | 4,17E-02 |
| | ENSG00000211677 | | immunoglobulin lambda constant 2 | 276,85 | 1,21 | 0,25 | 4,81 | 1,48E-06 | 7,48E-04 |
| CDCA7 | ENSG00000144354 | Q9BWT1 | cell division cycle associated 7 | 7,20 | 1,21 | 0,26 | 4,62 | 3,82E-06 | 1,66E-03 |
| CEP55 | ENSG00000138180 | Q53EZ4 | centrosomal protein 55 | 16,60 | 1,18 | 0,18 | 6,48 | 9,06E-11 | 3,48E-07 |
| MZB1 | ENSG00000170476 | Q8WU39 | marginal zone B and B1 cell specific protein | 17,48 | 1,17 | 0,29 | 4,03 | 5,66E-05 | 9,61E-03 |
| | ENSG00000209082 | | mitochondrially encoded tRNA-Leu (UUA/G) 1 | 224,75 | 1,16 | 0,32 | 3,64 | 2,70E-04 | 2,22E-02 |
| HMGB3 | ENSG00000029993 | O15347 | high mobility group box 3 | 5,79 | 1,16 | 0,24 | 4,87 | 1,12E-06 | 6,31E-04 |
| STMN1 | ENSG00000117632 | P16949 | stathmin 1 | 88,06 | 1,16 | 0,17 | 6,89 | 5,65E-12 | 4,20E-08 |
| TIMM10 | ENSG00000134809 | P62072 | translocase of inner mitochondrial membrane | 30,92 | 1,15 | 0,21 | 5,41 | 6,25E-08 | 6,35E-05 |
| KIF18B | ENSG00000186185 | Q86Y91 | kinesin family member 18B | 8,33 | 1,14 | 0,22 | 5,08 | 3,76E-07 | 2,78E-04 |
| | ENSG00000248587 | | GDNF antisense RNA 1 | 5,60 | 1,11 | 0,31 | 3,62 | 2,97E-04 | 2,34E-02 |
| | ENSG00000240087 | | ribosomal protein SA pseudogene 12 | 7,19 | 1,10 | 0,33 | 3,34 | 8,51E-04 | 4,27E-02 |
| | ENSG00000211679 | | immunoglobulin lambda constant 3 (Kern-Oz+ marker) | 261,71 | 1,10 | 0,26 | 4,23 | 2,30E-05 | 5,86E-03 |
| IGHG1 | ENSG00000211896 | P01857 | immunoglobulin heavy constant gamma 1 (G1m marker) | 40,14 | 1,08 | 0,33 | 3,32 | 8,97E-04 | 4,33E-02 |
| | ENSG00000231873 | | novel transcript | 38,11 | 1,08 | 0,25 | 4,31 | 1,60E-05 | 4,80E-03 |
| ANKRD62 | ENSG00000181626 | A6NC57 | ankyrin repeat domain 62 | 4,81 | 1,08 | 0,30 | 3,60 | 3,16E-04 | 2,41E-02 |
| TTK | ENSG00000112742 | P33981 | TTK protein kinase | 5,49 | 1,08 | 0,24 | 4,55 | 5,39E-06 | 2,05E-03 |
| RPS15A | ENSG00000134419 | P62244 | ribosomal protein S15a | 431,62 | 1,06 | 0,22 | 4,76 | 1,90E-06 | 9,35E-04 |
| LOC10050962( | ENSG00000259916 | A0A075B734 | novel member of the aquaporin (AQP) gene | 4,92 | 1,04 | 0,30 | 3,42 | 6,25E-04 | 3,59E-02 |
| CCNB1 | ENSG00000134057 | P14635 | cyclin B1 | 7,03 | 1,02 | 0,21 | 4,89 | 9,84E-07 | 5,72E-04 |
| KIFC1 | ENSG00000237649 | Q9BW19 | kinesin family member C1 | 15,06 | 1,02 | 0,18 | 5,73 | 1,02E-08 | 1,64E-05 |
| AURKB | ENSG00000178999 | Q96GD4 | aurora kinase B [Source:HGNC Symbol;Acc:HGNC:11390] | 10,45 | 1,02 | 0,25 | 4,11 | 4,01E-05 | 8,08E-03 |
| ASPM | ENSG00000066279 | Q8IZT6 | assembly factor for spindle microtubules | 34,50 | 1,00 | 0,19 | 5,41 | 6,28E-08 | 6,35E-05 |
| | ENSG00000226986 | | PRELID1 pseudogene 5 | 4,69 | 1,00 | 0,26 | 3,89 | 9,97E-05 | 1,27E-02 |
| CDC6 | ENSG00000094804 | Q99741 | cell division cycle 6 | 6,02 | 1,00 | 0,24 | 4,13 | 3,58E-05 | 7,47E-03 |
| SPAM1 | ENSG00000106304 | P38567 | sperm adhesion molecule 1 | 3,79 | 1,00 | 0,30 | 3,38 | 7,32E-04 | 3,92E-02 |
| | ENSG00000229526 | | keratin 16 pseudogene 4 [Source:HGNC Symbol;Acc:HGNC:37809] | 5,75 | 1,00 | 0,28 | 3,56 | 3,66E-04 | 2,63E-02 |
| | ENSG00000250777 | | novel transcript | 4,37 | 0,99 | 0,30 | 3,28 | 1,03E-03 | 4,57E-02 |
| H4C1 | ENSG00000273542 | P62805 | H4 clustered histone 12 | 5,59 | 0,99 | 0,30 | 3,29 | 1,00E-03 | 4,52E-02 |
| RRM2 | ENSG00000171848 | P31350 | ribonucleotide reductase regulatory subunit M2 | 55,17 | 0,99 | 0,14 | 6,87 | 6,57E-12 | 4,20E-08 |
| OTOF | ENSG00000115155 | Q9HC10 | otoferlin | 8,62 | 0,99 | 0,30 | 3,29 | 1,01E-03 | 4,52E-02 |
| | ENSG00000264350 | | small nuclear ribonucleoprotein polypeptide G pseudogene 2 | 8,70 | 0,99 | 0,28 | 3,51 | 4,44E-04 | 2,88E-02 |
| CKS1B | ENSG00000173207 | P61024 | CDC28 protein kinase regulatory subunit 1B | 20,83 | 0,98 | 0,17 | 5,68 | 1,31E-08 | 1,80E-05 |
| BTC | ENSG00000174808 | P35070 | betacellulin | 5,58 | 0,97 | 0,26 | 3,78 | 1,56E-04 | 1,65E-02 |
| GGH | ENSG00000137563 | Q92820 | gamma-glutamyl hydrolase | 6,40 | 0,97 | 0,24 | 4,01 | 5,96E-05 | 9,67E-03 |
| CENPF | ENSG00000117724 | P49454 | centromere protein F | 66,09 | 0,97 | 0,16 | 6,05 | 1,41E-09 | 3,39E-06 |
| | ENSG00000280594 | | BTG3 antisense RNA 1 | 3,67 | 0,97 | 0,27 | 3,57 | 3,56E-04 | 2,59E-02 |
| HNF1B | ENSG00000275410 | E0YMJ8 | HNF1 homeobox B | 6,34 | 0,95 | 0,28 | 3,37 | 7,60E-04 | 4,02E-02 |
| CDC20 | ENSG00000117399 | Q12834 | cell division cycle 20 | 5,34 | 0,94 | 0,28 | 3,38 | 7,32E-04 | 3,92E-02 |

| | | | | | | | | | |
|---|---|---|---|---|---|---|---|---|---|
| IGSF5 | ENSG00000183067 | Q9NSI5 | immunoglobulin superfamily member 5 | 7,45 | 0,94 | 0,26 | 3,57 | 3,51E-04 | 2,56E-02 |
| E2F7 | ENSG00000165891 | Q96AV8 | E2F transcription factor 7 | 9,50 | 0,94 | 0,20 | 4,62 | 3,90E-06 | 1,66E-03 |
| | ENSG00000253771 | | transmembrane phosphoinositide 3-phosphatase and tensin homolog 2 pseudogene | 10,40 | 0,93 | 0,23 | 3,97 | 7,12E-05 | 1,02E-02 |
| IL17D | ENSG00000172458 | Q8TAD2 | interleukin 17D | 3,89 | 0,92 | 0,29 | 3,23 | 1,22E-03 | 4,93E-02 |
| | ENSG00000285544 | | novel transcript | 5,67 | 0,92 | 0,24 | 3,81 | 1,42E-04 | 1,56E-02 |
| IGHA1 | ENSG00000211895 | P01876 | immunoglobulin heavy constant alpha 1 | 97,89 | 0,92 | 0,25 | 3,64 | 2,70E-04 | 2,22E-02 |
| | ENSG00000239039 | | small nucleolar RNA | 244,92 | 0,92 | 0,27 | 3,45 | 5,65E-04 | 3,36E-02 |
| MPP3 | ENSG00000161647 | Q13368 | membrane palmitoylated protein 3 | 4,86 | 0,90 | 0,26 | 3,40 | 6,73E-04 | 3,75E-02 |
| | ENSG00000232211 | | novel transcript | 4,52 | 0,90 | 0,27 | 3,33 | 8,57E-04 | 4,28E-02 |
| | ENSG00000244968 | | LIFR antisense RNA 1 | 7,09 | 0,90 | 0,24 | 3,70 | 2,12E-04 | 1,97E-02 |
| | ENSG00000285080 | | novel transcript | 44,89 | 0,88 | 0,24 | 3,66 | 2,53E-04 | 2,12E-02 |
| | ENSG00000259316 | | novel protein | 5,99 | 0,88 | 0,24 | 3,60 | 3,19E-04 | 2,41E-02 |
| GZMA | ENSG00000145649 | P12544 | granzyme A | 159,90 | 0,88 | 0,21 | 4,25 | 2,13E-05 | 5,67E-03 |
| HMMR | ENSG00000072571 | O75330 | hyaluronan mediated motility receptor | 8,60 | 0,87 | 0,22 | 3,92 | 8,68E-05 | 1,15E-02 |
| RPS21 | ENSG00000171858 | Q8WVC2 | ribosomal protein S21 | 1260,63 | 0,87 | 0,22 | 4,04 | 5,24E-05 | 9,23E-03 |
| RPL39 | ENSG00000198918 | P62891 | ribosomal protein L39 | 860,33 | 0,87 | 0,19 | 4,63 | 3,71E-06 | 1,66E-03 |
| FGF1 | ENSG00000113578 | P05230 | fibroblast growth factor 1 | 4,78 | 0,87 | 0,26 | 3,31 | 9,49E-04 | 4,39E-02 |
| CD52 | ENSG00000169442 | P31358 | CD52 molecule | 773,38 | 0,87 | 0,18 | 4,70 | 2,65E-06 | 1,27E-03 |
| NPM3 | ENSG00000107833 | O75607 | nucleophosmin/nucleoplasmin 3 | 98,49 | 0,87 | 0,24 | 3,62 | 2,99E-04 | 2,35E-02 |
| MCM10 | ENSG00000065328 | Q7L590 | minichromosome maintenance 10 replication initiation factor | 14,79 | 0,87 | 0,17 | 4,98 | 6,49E-07 | 4,02E-04 |
| SCRN2 | ENSG00000141295 | Q96FV2 | secernin 2 | 5,11 | 0,86 | 0,25 | 3,45 | 5,51E-04 | 3,30E-02 |
| CLEC19A | ENSG00000261210 | Q6UXS0 | C-type lectin domain containing 19A | 6,48 | 0,86 | 0,26 | 3,28 | 1,03E-03 | 4,57E-02 |
| MKI67 | ENSG00000148773 | P46013 | marker of proliferation Ki-67 | 84,79 | 0,86 | 0,19 | 4,51 | 6,48E-06 | 2,29E-03 |
| HINT1 | ENSG00000169567 | D6RC06 | histidine triad nucleotide binding protein 1 | 612,82 | 0,85 | 0,20 | 4,29 | 1,82E-05 | 5,22E-03 |
| | ENSG00000256686 | | novel transcript | 7,04 | 0,85 | 0,26 | 3,28 | 1,03E-03 | 4,57E-02 |
| | ENSG00000250166 | | novel transcript | 51,25 | 0,85 | 0,14 | 6,08 | 1,19E-09 | 3,26E-06 |
| SKA1 | ENSG00000154839 | Q96BD8 | spindle and kinetochore associated complex subunit 1 | 4,67 | 0,84 | 0,23 | 3,62 | 2,93E-04 | 2,34E-02 |
| RPL27 | ENSG00000131469 | A0A024R1V4 | ribosomal protein L27 | 416,72 | 0,83 | 0,21 | 3,98 | 6,92E-05 | 1,01E-02 |
| | ENSG00000274372 | | long intergenic non-protein coding RNA 2804 | 4,55 | 0,83 | 0,25 | 3,28 | 1,04E-03 | 4,60E-02 |
| ROMO1 | ENSG00000125995 | P60602 | reactive oxygen species modulator 1 | 148,18 | 0,83 | 0,22 | 3,83 | 1,28E-04 | 1,46E-02 |
| KCNK5 | ENSG00000164626 | O95279 | potassium two pore domain channel subfamily K member 5 | 5,97 | 0,83 | 0,24 | 3,48 | 5,04E-04 | 3,08E-02 |
| RPL22L1 | ENSG00000163584 | Q6P5R6 | ribosomal protein L22 like 1 | 23,99 | 0,83 | 0,24 | 3,39 | 6,89E-04 | 3,81E-02 |
| SULT1C4 | ENSG00000198075 | O75897 | sulfotransferase family 1C member 4 | 4,40 | 0,83 | 0,25 | 3,33 | 8,76E-04 | 4,31E-02 |
| EEF1B2 | ENSG00000114942 | P24534 | eukaryotic translation elongation factor 1 beta | 228,95 | 0,82 | 0,21 | 3,99 | 6,64E-05 | 9,86E-03 |
| ESCO2 | ENSG00000171320 | Q56NI9 | establishment of sister chromatid cohesion N-acetyltransferase 2 | 11,92 | 0,82 | 0,20 | 4,18 | 2,85E-05 | 6,56E-03 |
| TRAV30 | ENSG00000259092 | A0A087WSZ9 | T cell receptor alpha variable 30 | 10,57 | 0,81 | 0,24 | 3,41 | 6,61E-04 | 3,74E-02 |
| | ENSG00000255458 | | novel transcript | 10,42 | 0,81 | 0,22 | 3,70 | 2,17E-04 | 2,00E-02 |
| UQCRQ | ENSG00000164405 | O14949 | ubiquinol-cytochrome c reductase complex III subunit VII | 189,16 | 0,80 | 0,16 | 5,12 | 3,12E-07 | 2,50E-04 |
| RPA3 | ENSG00000106399 | P35244 | replication protein A3 | 35,48 | 0,80 | 0,20 | 4,01 | 6,03E-05 | 9,67E-03 |
| H4C1 | ENSG00000197238 | P62805 | H4 clustered histone 11 | 7,34 | 0,80 | 0,24 | 3,28 | 1,02E-03 | 4,55E-02 |
| CENPE | ENSG00000138778 | Q02224 | centromere protein E | 56,12 | 0,78 | 0,12 | 6,45 | 1,15E-10 | 3,66E-07 |
| TXNDC17 | ENSG00000129235 | Q9BRA2 | thioredoxin domain containing 17 | 42,48 | 0,77 | 0,17 | 4,55 | 5,45E-06 | 2,05E-03 |
| PTRHD1 | ENSG00000184924 | Q6GMV3 | peptidyl-tRNA hydrolase domain containing 1 | 25,42 | 0,77 | 0,19 | 4,08 | 4,49E-05 | 8,47E-03 |
| PAX2 | ENSG00000075891 | Q02962 | paired box 2 | 9,22 | 0,77 | 0,23 | 3,26 | 1,12E-03 | 4,75E-02 |
| EXO1 | ENSG00000174371 | Q9UQ84 | exonuclease 1 | 9,80 | 0,76 | 0,21 | 3,66 | 2,48E-04 | 2,11E-02 |
| CLSPN | ENSG00000092853 | Q9HAW4 | claspin | 44,19 | 0,76 | 0,15 | 5,23 | 1,72E-07 | 1,50E-04 |
| | ENSG00000262097 | | long intergenic non-protein coding RNA 2185 | 8,29 | 0,76 | 0,19 | 3,92 | 9,00E-05 | 1,17E-02 |
| | ENSG00000259882 | | rhophilin | 25,03 | 0,76 | 0,23 | 3,32 | 8,90E-04 | 4,33E-02 |
| | ENSG00000230342 | | FANCD2 pseudogene 2 | 8,14 | 0,75 | 0,23 | 3,25 | 1,14E-03 | 4,78E-02 |
| | ENSG00000229119 | | 60S acidic ribosomal protein (RPLP0) | 78,00 | 0,75 | 0,16 | 4,55 | 5,30E-06 | 2,05E-03 |
| RPS7 | ENSG00000171863 | P62081 | ribosomal protein S7 | 364,25 | 0,75 | 0,19 | 3,92 | 8,73E-05 | 1,15E-02 |
| | ENSG00000255282 | | Wilms tumor 1 associated protein pseudogene | 5,26 | 0,75 | 0,22 | 3,34 | 8,40E-04 | 4,26E-02 |
| SIT1 | ENSG00000137078 | Q9Y3P8 | signaling threshold regulating transmembrane adaptor 1 | 57,62 | 0,75 | 0,19 | 4,00 | 6,31E-05 | 9,85E-03 |
| RAB34 | ENSG00000109113 | P0DI83 | RAB34 | 13,95 | 0,74 | 0,18 | 4,12 | 3,74E-05 | 7,63E-03 |
| DBI | ENSG00000155368 | P07108 | diazepam binding inhibitor | 186,53 | 0,74 | 0,18 | 4,14 | 3,52E-05 | 7,42E-03 |
| IFI27L2 | ENSG00000119632 | Q9H2X8 | interferon alpha inducible protein 27 like 2 | 46,12 | 0,73 | 0,17 | 4,26 | 2,00E-05 | 5,49E-03 |
| KIF23 | ENSG00000137807 | Q02241 | kinesin family member 23 | 10,66 | 0,73 | 0,18 | 4,02 | 5,80E-05 | 9,67E-03 |
| COX7B | ENSG00000131174 | P24311 | cytochrome c oxidase subunit 7B | 41,78 | 0,73 | 0,20 | 3,70 | 2,13E-04 | 1,97E-02 |
| | ENSG00000268403 | | novel transcript | 6,76 | 0,73 | 0,21 | 3,47 | 5,12E-04 | 3,11E-02 |
| AYP1 | ENSG00000172922 | A0A024R5B3 | ribonuclease H2 subunit C | 62,04 | 0,72 | 0,17 | 4,27 | 1,94E-05 | 5,47E-03 |

| | | | | | | | | | |
|---|---|---|---|---|---|---|---|---|---|
| L1TD1 | ENSG00000240563 | Q5T7N2 | LINE1 type transposase domain containing 1 | 13,19 | 0,72 | 0,21 | 3,45 | 5,67E-04 | 3,36E-02 |
| RPS18 | ENSG00000231500 | P62269 | ribosomal protein S18 | 1401,14 | 0,72 | 0,20 | 3,66 | 2,51E-04 | 2,11E-02 |
| FBXO5 | ENSG00000112029 | Q9UKT4 | F-box protein 5 | 12,20 | 0,72 | 0,18 | 3,87 | 1,08E-04 | 1,32E-02 |
| PCNA | ENSG00000132646 | P12004 | proliferating cell nuclear antigen | 24,01 | 0,71 | 0,22 | 3,31 | 9,44E-04 | 4,39E-02 |
| RAN | ENSG00000132341 | B4DV51 | RAN | 347,28 | 0,71 | 0,19 | 3,77 | 1,60E-04 | 1,68E-02 |
| | ENSG00000254840 | | General transcription factor II-I (GTF2I) pseudogene | 9,79 | 0,70 | 0,19 | 3,63 | 2,89E-04 | 2,32E-02 |
| H1-2 | ENSG00000187837 | P16403 | H1.2 linker histone | 552,88 | 0,70 | 0,18 | 3,96 | 7,54E-05 | 1,06E-02 |
| CDCA2 | ENSG00000184661 | Q69YH5 | cell division cycle associated 2 | 13,52 | 0,70 | 0,19 | 3,63 | 2,79E-04 | 2,27E-02 |
| | ENSG00000255308 | | CSRP3 and E2F8 antisense RNA 1 | 11,56 | 0,70 | 0,19 | 3,61 | 3,11E-04 | 2,40E-02 |
| DTL | ENSG00000143476 | Q9NZJ0 | denticleless E3 ubiquitin protein ligase homolog | 29,27 | 0,70 | 0,15 | 4,81 | 1,47E-06 | 7,48E-04 |
| TSSK6 | ENSG00000178093 | A0A024R7Q5 | testis specific serine kinase 6 | 10,23 | 0,70 | 0,19 | 3,58 | 3,41E-04 | 2,52E-02 |
| TMEM213 | ENSG00000214128 | A2RRL7 | transmembrane protein 213 | 9,44 | 0,70 | 0,21 | 3,29 | 1,01E-03 | 4,52E-02 |
| RPL31 | ENSG00000071082 | P62899 | ribosomal protein L31 | 606,60 | 0,70 | 0,21 | 3,27 | 1,08E-03 | 4,66E-02 |
| TMEM258 | ENSG00000134825 | P61165 | transmembrane protein 258 | 198,64 | 0,69 | 0,16 | 4,44 | 9,15E-06 | 2,98E-03 |
| H4C1 | ENSG00000197061 | P62805 | H4 clustered histone 3 | 941,47 | 0,69 | 0,21 | 3,31 | 9,25E-04 | 4,39E-02 |
| C4orf48 | ENSG00000243449 | Q5BLP8 | chromosome 4 open reading frame 48 | 15,87 | 0,69 | 0,21 | 3,28 | 1,05E-03 | 4,61E-02 |
| SHCBP1 | ENSG00000171241 | Q8NEM2 | SHC binding and spindle associated 1 | 12,82 | 0,69 | 0,17 | 4,06 | 4,91E-05 | 9,13E-03 |
| CCNQ | ENSG00000262919 | Q8N1B3 | cyclin Q | 37,85 | 0,69 | 0,19 | 3,56 | 3,75E-04 | 2,66E-02 |
| | ENSG00000274979 | | novel transcript | 16,17 | 0,68 | 0,18 | 3,81 | 1,39E-04 | 1,55E-02 |
| RPL41 | ENSG00000229117 | P62945 | ribosomal protein L41 | 3867,40 | 0,68 | 0,17 | 3,99 | 6,49E-05 | 9,86E-03 |
| VAMP5 | ENSG00000168899 | O95183 | vesicle associated membrane protein 5 | 39,08 | 0,67 | 0,20 | 3,41 | 6,39E-04 | 3,65E-02 |
| MCM4 | ENSG00000104738 | P33991 | minichromosome maintenance complex component 4 | 18,58 | 0,67 | 0,17 | 4,01 | 6,11E-05 | 9,69E-03 |
| | ENSG00000257817 | | novel transcript | 22,61 | 0,67 | 0,20 | 3,31 | 9,38E-04 | 4,39E-02 |
| CHEK1 | ENSG00000149554 | O14757 | checkpoint kinase 1 | 19,06 | 0,67 | 0,15 | 4,57 | 4,95E-06 | 2,02E-03 |
| | ENSG00000226777 | | family with sequence similarity 30 member A | 37,79 | 0,66 | 0,19 | 3,52 | 4,39E-04 | 2,87E-02 |
| RPL35 | ENSG00000136942 | P42766 | ribosomal protein L35 | 201,40 | 0,66 | 0,17 | 3,79 | 1,54E-04 | 1,65E-02 |
| DLGAP5 | ENSG00000126787 | Q15398 | DLG associated protein 5 | 21,28 | 0,65 | 0,16 | 3,94 | 8,04E-05 | 1,09E-02 |
| RPS20 | ENSG00000008988 | P60866 | ribosomal protein S20 | 3959,06 | 0,65 | 0,15 | 4,38 | 1,19E-05 | 3,80E-03 |
| | ENSG00000203809 | | LIN28B antisense RNA 1 | 14,48 | 0,64 | 0,20 | 3,26 | 1,13E-03 | 4,78E-02 |
| COPS9 | ENSG00000172428 | Q8WXC6 | COP9 signalosome subunit 9 | 103,23 | 0,64 | 0,15 | 4,20 | 2,64E-05 | 6,32E-03 |
| XRCC2 | ENSG00000196584 | O43543 | X-ray repair cross complementing 2 | 15,56 | 0,64 | 0,16 | 4,05 | 5,16E-05 | 9,20E-03 |
| PDPN | ENSG00000162493 | Q86YL7 | podoplanin | 7,97 | 0,64 | 0,20 | 3,23 | 1,23E-03 | 4,93E-02 |
| NDUFB1 | ENSG00000183648 | O75438 | NADH:ubiquinone oxidoreductase subunit B1 | 272,08 | 0,64 | 0,17 | 3,69 | 2,23E-04 | 2,01E-02 |
| NCAPH | ENSG00000121152 | Q15003 | non-SMC condensin I complex subunit H | 23,35 | 0,64 | 0,15 | 4,22 | 2,44E-05 | 5,92E-03 |
| | ENSG00000246528 | | novel transcript | 14,09 | 0,64 | 0,16 | 3,89 | 1,02E-04 | 1,27E-02 |
| RPSA | ENSG00000168028 | P08865 | ribosomal protein SA | 489,37 | 0,63 | 0,17 | 3,77 | 1,66E-04 | 1,70E-02 |
| RFK | ENSG00000135002 | Q969G6 | riboflavin kinase [Source:HGNC Symbol;Acc:HGNC:30324] | 19,69 | 0,62 | 0,19 | 3,22 | 1,26E-03 | 4,94E-02 |
| NDUFS6 | ENSG00000145494 | O75380 | NADH:ubiquinone oxidoreductase subunit S6 | 264,47 | 0,62 | 0,13 | 4,62 | 3,84E-06 | 1,66E-03 |
| | ENSG00000136149 | | ribosomal protein L13a pseudogene 25 | 2038,88 | 0,62 | 0,17 | 3,72 | 1,98E-04 | 1,87E-02 |
| DPM3 | ENSG00000179085 | Q9P2X0 | dolichyl-phosphate mannosyltransferase subunit 3 | 67,77 | 0,61 | 0,16 | 3,89 | 1,02E-04 | 1,27E-02 |
| TIMM8B | ENSG00000150779 | Q9Y5J9 | translocase of inner mitochondrial membrane 8 homolog B | 19,74 | 0,61 | 0,19 | 3,27 | 1,06E-03 | 4,61E-02 |
| UQCR10 | ENSG00000184076 | Q9UDW1 | ubiquinol-cytochrome c reductase | 77,77 | 0,61 | 0,15 | 4,00 | 6,21E-05 | 9,77E-03 |
| GINS3 | ENSG00000181938 | Q9BRX5 | GINS complex subunit 3 | 31,48 | 0,61 | 0,17 | 3,56 | 3,64E-04 | 2,63E-02 |
| BOLA2 | ENSG00000169627 | Q9H3K6 | bolA family member 2B | 87,08 | 0,61 | 0,15 | 3,99 | 6,55E-05 | 9,86E-03 |
| PRELID3B | ENSG00000101166 | Q9Y3B1 | PRELI domain containing 3B | 13,05 | 0,60 | 0,18 | 3,30 | 9,50E-04 | 4,39E-02 |
| ATP5IF1 | ENSG00000130770 | Q9UII2 | ATP synthase inhibitory factor subunit 1 | 202,27 | 0,60 | 0,11 | 5,30 | 1,15E-07 | 1,06E-04 |
| COX7C | ENSG00000127184 | P15954 | cytochrome c oxidase subunit 7C | 442,34 | 0,60 | 0,18 | 3,34 | 8,41E-04 | 4,26E-02 |
| TPX2 | ENSG00000088325 | Q9ULW0 | TPX2 microtubule nucleation factor | 32,51 | 0,59 | 0,14 | 4,16 | 3,25E-05 | 7,05E-03 |
| TMEM70 | ENSG00000175606 | Q9BUB7 | transmembrane protein 70 | 25,26 | 0,59 | 0,17 | 3,44 | 5,73E-04 | 3,38E-02 |
| | ENSG00000240040 | | novel transcript | 49,13 | 0,59 | 0,17 | 3,40 | 6,71E-04 | 3,75E-02 |
| MRPL51 | ENSG00000111639 | Q4U2R6 | mitochondrial ribosomal protein L51 | 87,46 | 0,58 | 0,15 | 3,95 | 7,89E-05 | 1,08E-02 |
| CKAP2L | ENSG00000169607 | Q8IYA6 | cytoskeleton associated protein 2 like | 11,08 | 0,58 | 0,18 | 3,26 | 1,11E-03 | 4,75E-02 |
| | ENSG00000238063 | | long intergenic non-protein coding RNA 1685 | 14,83 | 0,58 | 0,18 | 3,31 | 9,49E-04 | 4,39E-02 |
| CYC1 | ENSG00000179091 | P08574 | cytochrome c1 | 33,62 | 0,58 | 0,15 | 3,78 | 1,56E-04 | 1,65E-02 |
| MYO5B | ENSG00000167306 | Q9ULV0 | myosin VB | 31,42 | 0,58 | 0,17 | 3,32 | 8,88E-04 | 4,33E-02 |
| UQCRB | ENSG00000156467 | B7Z2R2 | ubiquinol-cytochrome c reductase binding | 498,15 | 0,57 | 0,13 | 4,52 | 6,15E-06 | 2,27E-03 |
| RPS27A | ENSG00000143947 | P62979 | ribosomal protein S27a | 1370,84 | 0,57 | 0,16 | 3,55 | 3,84E-04 | 2,67E-02 |
| SDF2L1 | ENSG00000128228 | Q9HCN8 | stromal cell derived factor 2 like 1 | 87,53 | 0,57 | 0,17 | 3,27 | 1,06E-03 | 4,61E-02 |
| NDUFA6 | ENSG00000184983 | P56556 | NADH:ubiquinone oxidoreductase subunit A6 | 153,63 | 0,56 | 0,10 | 5,74 | 9,34E-09 | 1,63E-05 |
| SNRPD2 | ENSG00000125743 | P62316 | small nuclear ribonucleoprotein D2 polypeptide | 131,88 | 0,56 | 0,15 | 3,75 | 1,79E-04 | 1,77E-02 |

| | | | | | | | | | |
|---|---|---|---|---|---|---|---|---|---|
| TRAPPC2L | ENSG00000167515 | Q9UL33 | trafficking protein particle complex 2 like | 34,82 | 0,56 | 0,16 | 3,56 | 3,72E-04 | 2,66E-02 |
| POLQ | ENSG00000051341 | O75417 | DNA polymerase theta | 44,97 | 0,56 | 0,11 | 4,96 | 7,20E-07 | 4,32E-04 |
| | ENSG00000280383 | | novel transcript | 27,18 | 0,56 | 0,14 | 4,01 | 5,99E-05 | 9,67E-03 |
| MRPL54 | ENSG00000183617 | Q6P161 | mitochondrial ribosomal protein L54 | 28,00 | 0,55 | 0,14 | 3,94 | 8,15E-05 | 1,10E-02 |
| NDC80 | ENSG00000080986 | O14777 | NDC80 kinetochore complex component | 43,00 | 0,55 | 0,13 | 4,18 | 2,87E-05 | 6,56E-03 |
| NDUFS5 | ENSG00000168653 | O43920 | NADH:ubiquinone oxidoreductase subunit S5 | 181,57 | 0,55 | 0,14 | 3,82 | 1,32E-04 | 1,50E-02 |
| | ENSG00000236552 | | ribosomal protein L13a pseudogene 5 | 481,45 | 0,55 | 0,15 | 3,61 | 3,09E-04 | 2,40E-02 |
| ARHGEF33 | ENSG00000214694 | H7C1C9 | Rho guanine nucleotide exchange factor 33 | 12,02 | 0,55 | 0,17 | 3,27 | 1,08E-03 | 4,66E-02 |
| MELK | ENSG00000165304 | Q14680 | maternal embryonic leucine zipper kinase | 48,31 | 0,54 | 0,12 | 4,47 | 7,75E-06 | 2,61E-03 |
| GRIN2A | ENSG00000183454 | Q12879 | glutamate ionotropic receptor NMDA type subunit 2A | 44,71 | 0,54 | 0,16 | 3,29 | 9,97E-04 | 4,52E-02 |
| CIP2A | ENSG00000163507 | Q8TCG1 | cellular inhibitor of PP2A | 44,73 | 0,54 | 0,12 | 4,50 | 6,68E-06 | 2,29E-03 |
| COA3 | ENSG00000183978 | Q9Y2R0 | cytochrome c oxidase assembly factor 3 | 41,47 | 0,54 | 0,14 | 3,77 | 1,66E-04 | 1,70E-02 |
| SSR3 | ENSG00000114850 | Q9UNL2 | signal sequence receptor subunit 3 | 81,65 | 0,54 | 0,13 | 4,05 | 5,06E-05 | 9,16E-03 |
| MRPL43 | ENSG00000055950 | Q8N983 | mitochondrial ribosomal protein L43 | 19,65 | 0,54 | 0,17 | 3,22 | 1,28E-03 | 4,96E-02 |
| NTSR1 | ENSG00000101188 | P30989 | neurotensin receptor 1 | 27,87 | 0,53 | 0,16 | 3,36 | 7,70E-04 | 4,04E-02 |
| RPL37A | ENSG00000197756 | P61513 | ribosomal protein L37a | 7829,90 | 0,53 | 0,15 | 3,44 | 5,81E-04 | 3,41E-02 |
| CCDC34 | ENSG00000109881 | Q96HJ3 | coiled-coil domain containing 34 | 24,12 | 0,53 | 0,13 | 3,96 | 7,48E-05 | 1,06E-02 |
| | ENSG00000215424 | | MCM3AP antisense RNA 1 | 38,64 | 0,53 | 0,15 | 3,41 | 6,48E-04 | 3,69E-02 |
| DEPDC1B | ENSG00000035499 | Q8WUY9 | DEP domain containing 1B | 19,15 | 0,52 | 0,14 | 3,88 | 1,05E-04 | 1,29E-02 |
| RPL27A | ENSG00000166441 | P46776 | ribosomal protein L27a | 2490,46 | 0,52 | 0,14 | 3,82 | 1,33E-04 | 1,50E-02 |
| TMEM208 | ENSG00000168701 | Q9BTX3 | transmembrane protein 208 | 44,15 | 0,52 | 0,11 | 4,82 | 1,42E-06 | 7,48E-04 |

***Supplementary Table S4:*** ***Top 200 GO categories related to viral processes and host immune responses of DEGs during HZ episode versus those one year after HZ.***

| GO.ID | Term | Annotated | Significant | Expected | P-value |
|---|---|---|---|---|---|
| GO:0006614 | SRP-dependent cotranslational protein targeting to membrane | 91 | 35 | 4,11 | 5,90E-24 |
| GO:0006413 | translational initiation | 176 | 42 | 7,94 | 5,70E-22 |
| GO:0000184 | nuclear-transcribed mRNA catabolic process, nonsense-mediated decay | 109 | 36 | 4,92 | 5,80E-22 |
| GO:0019083 | viral transcription | 163 | 40 | 7,36 | 1,00E-20 |
| GO:0070125 | mitochondrial translational elongation | 70 | 22 | 3,16 | 2,00E-13 |
| GO:0070126 | mitochondrial translational termination | 73 | 22 | 3,29 | 5,30E-13 |
| GO:0002181 | cytoplasmic translation | 88 | 18 | 3,97 | 1,60E-09 |
| GO:0031145 | anaphase-promoting complex-dependent catabolic process | 77 | 17 | 3,47 | 1,40E-07 |
| GO:0051301 | cell division | 539 | 50 | 24,32 | 5,40E-07 |
| GO:0006364 | rRNA processing | 204 | 24 | 9,21 | 5,00E-05 |
| GO:0006123 | mitochondrial electron transport, cytochrome c to oxygen | 17 | 6 | 0,77 | 6,70E-05 |
| GO:0010972 | negative regulation of G2/M transition of mitotic cell cycle | 86 | 14 | 3,88 | 7,60E-05 |
| GO:0006120 | mitochondrial electron transport, NADH to ubiquinone | 47 | 10 | 2,12 | 8,50E-05 |
| GO:0006122 | mitochondrial electron transport, ubiquinol to cytochrome c | 12 | 5 | 0,54 | 1,10E-04 |
| GO:0002479 | antigen processing and presentation of exogenous peptide antigen via MHC class I, TAP-dependent | 65 | 11 | 2,93 | 1,40E-04 |
| GO:0042769 | DNA damage response, detection of DNA damage | 36 | 8 | 1,62 | 1,60E-04 |
| GO:0050852 | T cell receptor signaling pathway | 189 | 20 | 8,53 | 2,00E-04 |
| GO:0007052 | mitotic spindle organization | 111 | 14 | 5,01 | 2,40E-04 |
| GO:0032981 | mitochondrial respiratory chain complex I assembly | 58 | 10 | 2,62 | 2,40E-04 |
| GO:0007094 | mitotic spindle assembly checkpoint | 34 | 9 | 1,53 | 2,50E-04 |
| GO:0042776 | mitochondrial ATP synthesis coupled proton transport | 17 | 6 | 0,77 | 2,60E-04 |
| GO:0000209 | protein polyubiquitination | 312 | 29 | 14,08 | 2,70E-04 |
| GO:0050853 | B cell receptor signaling pathway | 106 | 14 | 4,78 | 2,90E-04 |
| GO:0000082 | G1/S transition of mitotic cell cycle | 231 | 29 | 10,42 | 3,10E-04 |
| GO:0006521 | regulation of cellular amino acid metabolic process | 52 | 10 | 2,35 | 3,50E-04 |
| GO:0061418 | regulation of transcription from RNA polymerase II promoter in response to hypoxia | 70 | 12 | 3,16 | 3,60E-04 |
| GO:0035722 | interleukin-12-mediated signaling pathway | 41 | 8 | 1,85 | 4,20E-04 |
| GO:0045039 | protein insertion into mitochondrial inner membrane | 9 | 4 | 0,41 | 4,30E-04 |

| | | | | | |
|---|---|---|---|---|---|
| GO:0000387 | spliceosomal snRNP assembly | 44 | 9 | 1,99 | 5,40E-04 |
| GO:0070498 | interleukin-1-mediated signaling pathway | 96 | 14 | 4,33 | 5,60E-04 |
| GO:0042274 | ribosomal small subunit biogenesis | 68 | 12 | 3,07 | 6,70E-04 |
| GO:0060337 | type I interferon signaling pathway | 78 | 10 | 3,52 | 9,40E-04 |
| GO:0051607 | defense response to virus | 222 | 22 | 10,02 | 9,50E-04 |
| GO:0045653 | negative regulation of megakaryocyte differentiation | 18 | 5 | 0,81 | 9,70E-04 |
| GO:0000028 | ribosomal small subunit assembly | 18 | 5 | 0,81 | 9,70E-04 |
| GO:0016032 | viral process | 859 | 95 | 38,76 | 1,02E-03 |
| GO:0038095 | Fc-epsilon receptor signaling pathway | 150 | 16 | 6,77 | 1,22E-03 |
| GO:0010499 | proteasomal ubiquitin-independent protein catabolic process | 19 | 5 | 0,86 | 1,26E-03 |
| GO:0031146 | SCF-dependent proteasomal ubiquitin-dependent protein catabolic process | 85 | 11 | 3,84 | 1,49E-03 |
| GO:0031936 | negative regulation of chromatin silencing | 12 | 4 | 0,54 | 1,52E-03 |
| GO:0034080 | CENP-A containing nucleosome assembly | 39 | 7 | 1,76 | 1,61E-03 |
| GO:0051444 | negative regulation of ubiquitin-protein transferase activity | 15 | 6 | 0,68 | 1,63E-03 |
| GO:0032020 | ISG15-protein conjugation | 6 | 3 | 0,27 | 1,65E-03 |
| GO:0006915 | apoptotic process | 1676 | 103 | 75,63 | 1,67E-03 |
| GO:0006260 | DNA replication | 255 | 30 | 11,51 | 1,73E-03 |
| GO:1901984 | negative regulation of protein acetylation | 20 | 4 | 0,90 | 2,03E-03 |
| GO:0099178 | regulation of retrograde trans-synaptic signaling by | 2 | 2 | 0,09 | 2,03E-03 |
| GO:1903094 | negative regulation of protein K48-linked deubiquitination | 2 | 2 | 0,09 | 2,03E-03 |
| GO:0021935 | cerebellar granule cell precursor tangential migration | 2 | 2 | 0,09 | 2,03E-03 |
| GO:2000157 | negative regulation of ubiquitin-specific protease activity | 2 | 2 | 0,09 | 2,03E-03 |
| GO:0045071 | negative regulation of viral genome replication | 45 | 7 | 2,03 | 2,31E-03 |
| GO:0000056 | ribosomal small subunit export from nucleus | 7 | 3 | 0,32 | 2,79E-03 |
| GO:0060267 | positive regulation of respiratory burst | 7 | 3 | 0,32 | 2,79E-03 |
| GO:0043983 | histone H4-K12 acetylation | 7 | 3 | 0,32 | 2,79E-03 |
| GO:0000027 | ribosomal large subunit assembly | 23 | 5 | 1,04 | 3,15E-03 |
| GO:0038061 | NIK/NF-kappaB signaling | 151 | 12 | 6,81 | 3,73E-03 |
| GO:1901796 | regulation of signal transduction by p53 class mediator | 157 | 16 | 7,08 | 3,91E-03 |
| GO:1902036 | regulation of hematopoietic stem cell differentiation | 70 | 9 | 3,16 | 4,08E-03 |
| GO:0015014 | heparan sulfate proteoglycan biosynthetic process, polysaccharide chain biosynthetic process | 8 | 3 | 0,36 | 4,32E-03 |
| GO:0038096 | Fc-gamma receptor signaling pathway involved in phagocytosis | 119 | 13 | 5,37 | 4,59E-03 |
| GO:0007057 | spindle assembly involved in female meiosis I | 3 | 2 | 0,14 | 5,92E-03 |
| GO:0007089 | traversing start control point of mitotic cell cycle | 3 | 2 | 0,14 | 5,92E-03 |
| GO:0043335 | protein unfolding | 3 | 2 | 0,14 | 5,92E-03 |
| GO:1902445 | regulation of mitochondrial membrane permeability involved in programmed necrotic cell death | 3 | 2 | 0,14 | 5,92E-03 |
| GO:2000435 | negative regulation of protein neddylation | 3 | 2 | 0,14 | 5,92E-03 |
| GO:0043988 | histone H3-S28 phosphorylation | 3 | 2 | 0,14 | 5,92E-03 |
| GO:0001556 | oocyte maturation | 21 | 5 | 0,95 | 6,09E-03 |
| GO:1904667 | negative regulation of ubiquitin protein ligase activity | 9 | 3 | 0,41 | 6,27E-03 |
| GO:0010273 | detoxification of copper ion | 9 | 3 | 0,41 | 6,27E-03 |
| GO:0000245 | spliceosomal complex assembly | 64 | 7 | 2,89 | 6,50E-03 |
| GO:0006457 | protein folding | 201 | 20 | 9,07 | 6,67E-03 |
| GO:0070317 | negative regulation of G0 to G1 transition | 39 | 6 | 1,76 | 7,56E-03 |
| GO:0043011 | myeloid dendritic cell differentiation | 18 | 4 | 0,81 | 7,58E-03 |
| GO:0071294 | cellular response to zinc ion | 18 | 4 | 0,81 | 7,58E-03 |
| GO:0006977 | DNA damage response, signal transduction by p53 class mediator resulting in cell cycle arrest | 51 | 7 | 2,30 | 7,60E-03 |
| GO:0042407 | cristae formation | 28 | 5 | 1,26 | 7,64E-03 |
| GO:0002223 | stimulatory C-type lectin receptor signaling pathway | 108 | 11 | 4,87 | 9,49E-03 |
| GO:0071897 | DNA biosynthetic process | 170 | 18 | 7,67 | 1,08E-02 |

| | | | | | |
|---|---|---|---|---|---|
| GO:0048025 | negative regulation of mRNA splicing, via spliceosome | 20 | 4 | 0,90 | 1,12E-02 |
| GO:0010332 | response to gamma radiation | 46 | 5 | 2,08 | 1,12E-02 |
| GO:0051383 | kinetochore organization | 18 | 5 | 0,81 | 1,14E-02 |
| GO:0046931 | pore complex assembly | 20 | 4 | 0,90 | 1,15E-02 |
| GO:1902255 | positive regulation of intrinsic apoptotic signaling pathway by p53 class mediator | 4 | 2 | 0,18 | 1,15E-02 |
| GO:1905448 | positive regulation of mitochondrial ATP synthesis coupled electron transport | 4 | 2 | 0,18 | 1,15E-02 |
| GO:0051481 | negative regulation of cytosolic calcium ion concentration | 11 | 3 | 0,50 | 1,15E-02 |
| GO:0031115 | negative regulation of microtubule polymerization | 11 | 3 | 0,50 | 1,15E-02 |
| GO:0000727 | double-strand break repair via break-induced replication | 11 | 3 | 0,50 | 1,15E-02 |
| GO:0030210 | heparin biosynthetic process | 11 | 3 | 0,50 | 1,15E-02 |
| GO:0043488 | regulation of mRNA stability | 175 | 14 | 7,90 | 1,16E-02 |
| GO:0071276 | cellular response to cadmium ion | 31 | 5 | 1,40 | 1,18E-02 |
| GO:0016579 | protein deubiquitination | 247 | 22 | 11,15 | 1,24E-02 |
| GO:0018105 | peptidyl-serine phosphorylation | 283 | 25 | 12,77 | 1,33E-02 |
| GO:0050773 | regulation of dendrite development | 93 | 10 | 4,20 | 1,46E-02 |
| GO:0001682 | tRNA 5'-leader removal | 12 | 3 | 0,54 | 1,48E-02 |
| GO:0007059 | chromosome segregation | 299 | 32 | 13,49 | 1,51E-02 |
| GO:0006270 | DNA replication initiation | 39 | 6 | 1,76 | 1,53E-02 |
| GO:0060071 | Wnt signaling pathway, planar cell polarity pathway | 101 | 10 | 4,56 | 1,57E-02 |
| GO:0006955 | immune response | 1992 | 119 | 89,89 | 1,66E-02 |
| GO:0019985 | translesion synthesis | 41 | 6 | 1,85 | 1,85E-02 |
| GO:0045541 | negative regulation of cholesterol biosynthetic process | 5 | 2 | 0,23 | 1,86E-02 |
| GO:0071035 | nuclear polyadenylation-dependent rRNA catabolic process | 5 | 2 | 0,23 | 1,86E-02 |
| GO:0071038 | nuclear polyadenylation-dependent tRNA catabolic process | 5 | 2 | 0,23 | 1,86E-02 |
| GO:0032466 | negative regulation of cytokinesis | 5 | 2 | 0,23 | 1,86E-02 |
| GO:0008635 | activation of cysteine-type endopeptidase activity involved in apoptotic process by cytochrome c | 5 | 2 | 0,23 | 1,86E-02 |
| GO:0019919 | peptidyl-arginine methylation, to asymmetrical-dimethyl | 5 | 2 | 0,23 | 1,86E-02 |
| GO:0071947 | protein deubiquitination involved in ubiquitin-dependent protein catabolic process | 5 | 2 | 0,23 | 1,86E-02 |
| GO:0060391 | positive regulation of SMAD protein signal transduction | 13 | 3 | 0,59 | 1,86E-02 |
| GO:0045059 | positive thymic T cell selection | 13 | 3 | 0,59 | 1,86E-02 |
| GO:0006910 | phagocytosis, recognition | 76 | 8 | 3,43 | 2,09E-02 |
| GO:0006974 | cellular response to DNA damage stimulus | 795 | 67 | 35,88 | 2,18E-02 |
| GO:0019886 | antigen processing and presentation of exogenous peptide antigen via MHC class II | 92 | 9 | 4,15 | 2,29E-02 |
| GO:0006271 | DNA strand elongation involved in DNA replication | 18 | 4 | 0,81 | 2,29E-02 |
| GO:0034501 | protein localization to kinetochore | 18 | 4 | 0,81 | 2,29E-02 |
| GO:0001732 | formation of cytoplasmic translation initiation complex | 14 | 3 | 0,63 | 2,30E-02 |
| GO:0051382 | kinetochore assembly | 14 | 3 | 0,63 | 2,30E-02 |
| GO:1903214 | regulation of protein targeting to mitochondrion | 41 | 7 | 1,85 | 2,68E-02 |
| GO:0046825 | regulation of protein export from nucleus | 30 | 5 | 1,35 | 2,69E-02 |
| GO:1905618 | positive regulation of miRNA mediated inhibition of | 6 | 2 | 0,27 | 2,70E-02 |
| GO:0000055 | ribosomal large subunit export from nucleus | 6 | 2 | 0,27 | 2,70E-02 |
| GO:1902177 | positive regulation of oxidative stress-induced intrinsic apoptotic signaling pathway | 6 | 2 | 0,27 | 2,70E-02 |
| GO:1903608 | protein localization to cytoplasmic stress granule | 6 | 2 | 0,27 | 2,70E-02 |
| GO:2000171 | negative regulation of dendrite development | 6 | 2 | 0,27 | 2,70E-02 |
| GO:0043985 | histone H4-R3 methylation | 6 | 2 | 0,27 | 2,70E-02 |
| GO:0090267 | positive regulation of mitotic cell cycle spindle assembly | 6 | 2 | 0,27 | 2,70E-02 |
| GO:0042273 | ribosomal large subunit biogenesis | 62 | 12 | 2,80 | 2,70E-02 |
| GO:0060828 | regulation of canonical Wnt signaling pathway | 263 | 17 | 11,87 | 2,75E-02 |
| GO:0016584 | nucleosome positioning | 15 | 3 | 0,68 | 2,77E-02 |
| GO:0072359 | circulatory system development | 957 | 30 | 43,19 | 2,82E-02 |

| | | | | | |
|---|---|---|---|---|---|
| GO:0042981 | regulation of apoptotic process | 1272 | 73 | 57,40 | 2,86E-02 |
| GO:0006958 | complement activation, classical pathway | 112 | 10 | 5,05 | 2,99E-02 |
| GO:0006367 | transcription initiation from RNA polymerase II promoter | 173 | 13 | 7,81 | 3,01E-02 |
| GO:0036297 | interstrand cross-link repair | 53 | 6 | 2,39 | 3,14E-02 |
| GO:0002230 | positive regulation of defense response to virus by host | 27 | 4 | 1,22 | 3,16E-02 |
| GO:0051170 | import into nucleus | 148 | 12 | 6,68 | 3,27E-02 |
| GO:0030261 | chromosome condensation | 40 | 7 | 1,81 | 3,27E-02 |
| GO:0000186 | activation of MAPKK activity | 50 | 6 | 2,26 | 3,28E-02 |
| GO:0007569 | cell aging | 94 | 7 | 4,24 | 3,30E-02 |
| GO:0043981 | histone H4-K5 acetylation | 16 | 3 | 0,72 | 3,30E-02 |
| GO:0043982 | histone H4-K8 acetylation | 16 | 3 | 0,72 | 3,30E-02 |
| GO:0008283 | cell population proliferation | 1664 | 78 | 75,09 | 3,51E-02 |
| GO:0045737 | positive regulation of cyclin-dependent protein serine/threonine kinase activity | 28 | 4 | 1,26 | 3,56E-02 |
| GO:0030071 | regulation of mitotic metaphase/anaphase transition | 55 | 13 | 2,48 | 3,59E-02 |
| GO:0010324 | membrane invagination | 119 | 12 | 5,37 | 3,62E-02 |
| GO:0046596 | regulation of viral entry into host cell | 33 | 4 | 1,49 | 3,66E-02 |
| GO:0060372 | regulation of atrial cardiac muscle cell membrane | 7 | 2 | 0,32 | 3,67E-02 |
| GO:0099527 | postsynapse to nucleus signaling pathway | 7 | 2 | 0,32 | 3,67E-02 |
| GO:0060586 | multicellular organismal iron ion homeostasis | 7 | 2 | 0,32 | 3,67E-02 |
| GO:0002317 | plasma cell differentiation | 7 | 2 | 0,32 | 3,67E-02 |
| GO:1901165 | positive regulation of trophoblast cell migration | 7 | 2 | 0,32 | 3,67E-02 |
| GO:0019064 | fusion of virus membrane with host plasma membrane | 7 | 2 | 0,32 | 3,67E-02 |
| GO:0051899 | membrane depolarization | 76 | 9 | 3,43 | 3,83E-02 |
| GO:0033617 | mitochondrial cytochrome c oxidase assembly | 17 | 3 | 0,77 | 3,88E-02 |
| GO:0006335 | DNA replication-dependent nucleosome assembly | 29 | 4 | 1,31 | 3,99E-02 |
| GO:0050871 | positive regulation of B cell activation | 119 | 10 | 5,37 | 3,99E-02 |
| GO:0090263 | positive regulation of canonical Wnt signaling pathway | 136 | 12 | 6,14 | 4,16E-02 |
| GO:0006911 | phagocytosis, engulfment | 103 | 9 | 4,65 | 4,30E-02 |
| GO:0033045 | regulation of sister chromatid segregation | 64 | 15 | 2,89 | 4,44E-02 |
| GO:0045648 | positive regulation of erythrocyte differentiation | 30 | 4 | 1,35 | 4,44E-02 |
| GO:0098586 | cellular response to virus | 58 | 5 | 2,62 | 4,45E-02 |
| GO:2000816 | negative regulation of mitotic sister chromatid separation | 37 | 11 | 1,67 | 4,46E-02 |
| GO:0038127 | ERBB signaling pathway | 137 | 9 | 6,18 | 4,50E-02 |
| GO:0035970 | peptidyl-threonine dephosphorylation | 18 | 3 | 0,81 | 4,50E-02 |
| GO:0030150 | protein import into mitochondrial matrix | 18 | 3 | 0,81 | 4,50E-02 |
| GO:0048026 | positive regulation of mRNA splicing, via spliceosome | 18 | 3 | 0,81 | 4,50E-02 |
| GO:0099607 | lateral attachment of mitotic spindle microtubules to | 2 | 2 | 0,09 | 4,51E-02 |
| GO:0043921 | modulation by host of viral transcription | 2 | 2 | 0,09 | 4,51E-02 |
| GO:1904044 | response to aldosterone | 4 | 2 | 0,18 | 4,51E-02 |
| GO:0036344 | platelet morphogenesis | 20 | 2 | 0,90 | 4,51E-02 |
| GO:0150116 | regulation of cell-substrate junction organization | 67 | 4 | 3,02 | 4,51E-02 |
| GO:0036333 | hepatocyte homeostasis | 1 | 1 | 0,05 | 4,51E-02 |
| GO:0036339 | lymphocyte adhesion to endothelial cell of high endothelial | 1 | 1 | 0,05 | 4,51E-02 |
| GO:2000661 | positive regulation of interleukin-1-mediated signaling | 1 | 1 | 0,05 | 4,51E-02 |
| GO:0061871 | negative regulation of hepatic stellate cell migration | 1 | 1 | 0,05 | 4,51E-02 |
| GO:0036471 | cellular response to glyoxal | 1 | 1 | 0,05 | 4,51E-02 |
| GO:0002939 | tRNA N1-guanine methylation | 1 | 1 | 0,05 | 4,51E-02 |
| GO:0060058 | positive regulation of apoptotic process involved in mammary gland involution | 1 | 1 | 0,05 | 4,51E-02 |
| GO:0090668 | endothelial cell chemotaxis to vascular endothelial growth | 1 | 1 | 0,05 | 4,51E-02 |
| GO:0090625 | mRNA cleavage involved in gene silencing by siRNA | 1 | 1 | 0,05 | 4,51E-02 |

| | | | | | |
|---|---|---|---|---|---|
| GO:1900036 | positive regulation of cellular response to heat | 1 | 1 | 0,05 | 4,51E-02 |
| GO:1901622 | positive regulation of smoothened signaling pathway involved in dorsal/ventral neural tube patterning | 1 | 1 | 0,05 | 4,51E-02 |
| GO:0071040 | nuclear polyadenylation-dependent antisense transcript catabolic process | 1 | 1 | 0,05 | 4,51E-02 |
| GO:0071045 | nuclear histone mRNA catabolic process | 1 | 1 | 0,05 | 4,51E-02 |
| GO:0071036 | nuclear polyadenylation-dependent snoRNA catabolic process | 1 | 1 | 0,05 | 4,51E-02 |
| GO:0071037 | nuclear polyadenylation-dependent snRNA catabolic process | 1 | 1 | 0,05 | 4,51E-02 |
| GO:0071039 | nuclear polyadenylation-dependent CUT catabolic process | 1 | 1 | 0,05 | 4,51E-02 |
| GO:0021539 | subthalamus development | 1 | 1 | 0,05 | 4,51E-02 |
| GO:1903096 | protein localization to meiotic spindle midzone | 1 | 1 | 0,05 | 4,51E-02 |
| GO:1903060 | negative regulation of protein lipidation | 1 | 1 | 0,05 | 4,51E-02 |
| GO:0045975 | positive regulation of translation, ncRNA-mediated | 1 | 1 | 0,05 | 4,51E-02 |
| GO:1903073 | negative regulation of death-inducing signaling complex assembly | 1 | 1 | 0,05 | 4,51E-02 |
| GO:1990968 | modulation by host of RNA binding by virus | 1 | 1 | 0,05 | 4,51E-02 |
| GO:1990969 | modulation by host of viral RNA-binding transcription factor activity | 1 | 1 | 0,05 | 4,51E-02 |
| GO:0106045 | guanine deglycation, methylglyoxal removal | 1 | 1 | 0,05 | 4,51E-02 |
| GO:0106046 | guanine deglycation, glyoxal removal | 1 | 1 | 0,05 | 4,51E-02 |
| GO:1904291 | positive regulation of mitotic DNA damage checkpoint | 1 | 1 | 0,05 | 4,51E-02 |
| GO:0061974 | perichondral bone morphogenesis | 1 | 1 | 0,05 | 4,51E-02 |
| GO:0071386 | cellular response to corticosterone stimulus | 1 | 1 | 0,05 | 4,51E-02 |
| GO:0072707 | cellular response to sodium dodecyl sulfate | 1 | 1 | 0,05 | 4,51E-02 |
| GO:0061910 | autophagosome-endosome fusion | 1 | 1 | 0,05 | 4,51E-02 |
| GO:0086024 | adenylate cyclase-activating adrenergic receptor signaling pathway involved in positive regulation of heart rate | 1 | 1 | 0,05 | 4,51E-02 |

***Supplementary Table S5: Top 200 GO categories related to viral processes and host immune responses of DEGs during HZ episode compared to controls.***

| GO.ID | Term | Annotated | Significant | Expected | P-value |
|---|---|---|---|---|---|
| GO:0006614 | SRP-dependent cotranslational protein targeting to membrane | 91 | 26 | 2,24 | 5,90E-21 |
| GO:0006413 | translational initiation | 176 | 28 | 4,33 | 1,80E-19 |
| GO:0000184 | nuclear-transcribed mRNA catabolic process, nonsense-mediated decay | 109 | 26 | 2,68 | 8,90E-19 |
| GO:0019083 | viral transcription | 163 | 26 | 4,01 | 7,10E-15 |
| GO:0002181 | cytoplasmic translation | 88 | 14 | 2,17 | 1,50E-09 |
| GO:0070125 | mitochondrial translational elongation | 70 | 13 | 1,72 | 1,30E-08 |
| GO:0007052 | mitotic spindle organization | 111 | 18 | 2,73 | 1,60E-08 |
| GO:0070126 | mitochondrial translational termination | 73 | 13 | 1,80 | 2,20E-08 |
| GO:0051301 | cell division | 539 | 41 | 13,26 | 3,20E-08 |
| GO:0006122 | mitochondrial electron transport, ubiquinol to cytochrome c | 12 | 6 | 0,30 | 1,70E-07 |
| GO:0031145 | anaphase-promoting complex-dependent catabolic process | 77 | 12 | 1,89 | 1,80E-06 |
| GO:0007094 | mitotic spindle assembly checkpoint | 34 | 8 | 0,84 | 8,30E-06 |
| GO:0006260 | DNA replication | 255 | 24 | 6,28 | 1,80E-05 |
| GO:0034080 | CENP-A containing nucleosome assembly | 39 | 7 | 0,96 | 4,00E-05 |
| GO:0006123 | mitochondrial electron transport, cytochrome c to oxygen | 17 | 5 | 0,42 | 4,30E-05 |
| GO:1905448 | positive regulation of mitochondrial ATP synthesis coupled electron transport | 4 | 3 | 0,10 | 5,80E-05 |
| GO:0032981 | mitochondrial respiratory chain complex I assembly | 58 | 8 | 1,43 | 8,20E-05 |
| GO:0000083 | regulation of transcription involved in G1/S transition of mitotic cell cycle | 34 | 7 | 0,84 | 9,20E-05 |

| | | | | | |
|---|---|---|---|---|---|
| GO:1901796 | regulation of signal transduction by p53 class mediator | 157 | 12 | 3,86 | 2,30E-04 |
| GO:0034501 | protein localization to kinetochore | 18 | 5 | 0,44 | 2,90E-04 |
| GO:0051382 | kinetochore assembly | 14 | 4 | 0,34 | 3,00E-04 |
| GO:0000082 | G1/S transition of mitotic cell cycle | 231 | 23 | 5,68 | 3,10E-04 |
| GO:0070317 | negative regulation of G0 to G1 transition | 39 | 6 | 0,96 | 3,50E-04 |
| GO:0010971 | positive regulation of G2/M transition of mitotic cell cycle | 27 | 5 | 0,66 | 4,50E-04 |
| GO:0006120 | mitochondrial electron transport, NADH to ubiquinone | 47 | 7 | 1,16 | 5,20E-04 |
| GO:0061188 | negative regulation of ribosomal DNA heterochromatin assembly | 2 | 2 | 0,05 | 6,00E-04 |
| GO:0099178 | regulation of retrograde trans-synaptic signaling by endocanabinoid | 2 | 2 | 0,05 | 6,00E-04 |
| GO:0007059 | chromosome segregation | 299 | 32 | 7,36 | 6,50E-04 |
| GO:0051256 | mitotic spindle midzone assembly | 8 | 3 | 0,20 | 7,60E-04 |
| GO:0090091 | positive regulation of extracellular matrix disassembly | 8 | 3 | 0,20 | 7,60E-04 |
| GO:0010499 | proteasomal ubiquitin-independent protein catabolic process | 19 | 4 | 0,47 | 1,04E-03 |
| GO:0045039 | protein insertion into mitochondrial inner membrane | 9 | 3 | 0,22 | 1,11E-03 |
| GO:0042274 | ribosomal small subunit biogenesis | 68 | 9 | 1,67 | 1,51E-03 |
| GO:0006977 | DNA damage response, signal transduction by p53 class mediator resulting in cell cycle arrest | 51 | 6 | 1,26 | 1,51E-03 |
| GO:0000460 | maturation of 5.8S rRNA | 32 | 4 | 0,79 | 1,55E-03 |
| GO:0042769 | DNA damage response, detection of DNA damage | 36 | 5 | 0,89 | 1,77E-03 |
| GO:0007057 | spindle assembly involved in female meiosis I | 3 | 2 | 0,07 | 1,78E-03 |
| GO:0016579 | protein deubiquitination | 247 | 16 | 6,08 | 2,30E-03 |
| GO:0000387 | spliceosomal snRNP assembly | 44 | 5 | 1,08 | 2,58E-03 |
| GO:0007019 | microtubule depolymerization | 41 | 6 | 1,01 | 3,46E-03 |
| GO:0007079 | mitotic chromosome movement towards spindle pole | 4 | 2 | 0,10 | 3,51E-03 |
| GO:0097681 | double-strand break repair via alternative nonhomologous end joining | 4 | 2 | 0,10 | 3,51E-03 |
| GO:1903690 | negative regulation of wound healing, spreading of epidermal cells | 4 | 2 | 0,10 | 3,51E-03 |
| GO:0016233 | telomere capping | 51 | 5 | 1,26 | 4,61E-03 |
| GO:0045737 | positive regulation of cyclin-dependent protein serine/threonine kinase activity | 28 | 4 | 0,69 | 4,63E-03 |
| GO:0000028 | ribosomal small subunit assembly | 18 | 4 | 0,44 | 5,36E-03 |
| GO:0040001 | establishment of mitotic spindle localization | 32 | 4 | 0,79 | 5,71E-03 |
| GO:0015990 | electron transport coupled proton transport | 5 | 2 | 0,12 | 5,75E-03 |
| GO:0032466 | negative regulation of cytokinesis | 5 | 2 | 0,12 | 5,75E-03 |
| GO:0010626 | negative regulation of Schwann cell proliferation | 5 | 2 | 0,12 | 5,75E-03 |
| GO:1904261 | positive regulation of basement membrane assembly involved in embryonic body morphogenesis | 5 | 2 | 0,12 | 5,75E-03 |
| GO:0051170 | import into nucleus | 148 | 5 | 3,64 | 6,58E-03 |
| GO:0001556 | oocyte maturation | 21 | 4 | 0,52 | 7,73E-03 |
| GO:0042776 | mitochondrial ATP synthesis coupled proton transport | 17 | 3 | 0,42 | 7,78E-03 |
| GO:0010972 | negative regulation of G2/M transition of mitotic cell cycle | 86 | 10 | 2,12 | 7,80E-03 |
| GO:0006270 | DNA replication initiation | 39 | 6 | 0,96 | 8,26E-03 |
| GO:0032435 | negative regulation of proteasomal ubiquitin-dependent protein catabolic process | 33 | 4 | 0,81 | 8,41E-03 |
| GO:1902975 | mitotic DNA replication initiation | 6 | 2 | 0,15 | 8,49E-03 |
| GO:1990379 | lipid transport across blood-brain barrier | 6 | 2 | 0,15 | 8,49E-03 |
| GO:1905618 | positive regulation of miRNA mediated inhibition of translation | 6 | 2 | 0,15 | 8,49E-03 |
| GO:0033864 | positive regulation of NAD(P)H oxidase activity | 6 | 2 | 0,15 | 8,49E-03 |
| GO:0000055 | ribosomal large subunit export from nucleus | 6 | 2 | 0,15 | 8,49E-03 |
| GO:0045542 | positive regulation of cholesterol biosynthetic process | 6 | 2 | 0,15 | 8,49E-03 |
| GO:0006289 | nucleotide-excision repair | 98 | 7 | 2,41 | 9,03E-03 |
| GO:0045653 | negative regulation of megakaryocyte differentiation | 18 | 3 | 0,44 | 9,17E-03 |
| GO:0090307 | mitotic spindle assembly | 61 | 8 | 1,50 | 9,30E-03 |
| GO:0030071 | regulation of mitotic metaphase/anaphase transition | 55 | 12 | 1,35 | 1,12E-02 |
| GO:0031023 | microtubule organizing center organization | 125 | 12 | 3,08 | 1,13E-02 |

| | | | | | |
|---|---|---|---|---|---|
| GO:0000056 | ribosomal small subunit export from nucleus | 7 | 2 | 0,17 | 1,17E-02 |
| GO:1901165 | positive regulation of trophoblast cell migration | 7 | 2 | 0,17 | 1,17E-02 |
| GO:0071897 | DNA biosynthetic process | 170 | 9 | 4,18 | 1,22E-02 |
| GO:0006911 | phagocytosis, engulfment | 103 | 7 | 2,53 | 1,36E-02 |
| GO:0042276 | error-prone translesion synthesis | 21 | 3 | 0,52 | 1,42E-02 |
| GO:0051983 | regulation of chromosome segregation | 80 | 15 | 1,97 | 1,45E-02 |
| GO:0006281 | DNA repair | 522 | 27 | 12,85 | 1,51E-02 |
| GO:0051298 | centrosome duplication | 63 | 5 | 1,55 | 1,52E-02 |
| GO:0033314 | mitotic DNA replication checkpoint | 8 | 2 | 0,20 | 1,53E-02 |
| GO:0042428 | serotonin metabolic process | 8 | 2 | 0,20 | 1,53E-02 |
| GO:0070314 | G1 to G0 transition | 8 | 2 | 0,20 | 1,53E-02 |
| GO:0032201 | telomere maintenance via semi-conservative replication | 24 | 4 | 0,59 | 1,60E-02 |
| GO:0070987 | error-free translesion synthesis | 22 | 3 | 0,54 | 1,61E-02 |
| GO:0006297 | nucleotide-excision repair, DNA gap filling | 22 | 3 | 0,54 | 1,61E-02 |
| GO:0048511 | rhythmic process | 268 | 12 | 6,60 | 1,78E-02 |
| GO:0035722 | interleukin-12-mediated signaling pathway | 41 | 4 | 1,01 | 1,79E-02 |
| GO:1903599 | positive regulation of autophagy of mitochondrion | 9 | 2 | 0,22 | 1,94E-02 |
| GO:1904667 | negative regulation of ubiquitin protein ligase activity | 9 | 2 | 0,22 | 1,94E-02 |
| GO:0007026 | negative regulation of microtubule depolymerization | 24 | 3 | 0,59 | 2,04E-02 |
| GO:0048146 | positive regulation of fibroblast proliferation | 43 | 4 | 1,06 | 2,09E-02 |
| GO:0016032 | viral process | 859 | 51 | 21,14 | 2,10E-02 |
| GO:0007018 | microtubule-based movement | 322 | 8 | 7,92 | 2,15E-02 |
| GO:0002479 | antigen processing and presentation of exogenous peptide antigen via MHC class I, TAP-dependent | 65 | 5 | 1,60 | 2,18E-02 |
| GO:0042407 | cristae formation | 28 | 4 | 0,69 | 2,26E-02 |
| GO:0006910 | phagocytosis, recognition | 76 | 6 | 1,87 | 2,29E-02 |
| GO:0072321 | chaperone-mediated protein transport | 10 | 2 | 0,25 | 2,39E-02 |
| GO:2000816 | negative regulation of mitotic sister chromatid separation | 37 | 10 | 0,91 | 2,41E-02 |
| GO:0072431 | signal transduction involved in mitotic G1 DNA damage checkpoint | 52 | 7 | 1,28 | 2,43E-02 |
| GO:0099607 | lateral attachment of mitotic spindle microtubules to kinetochore | 2 | 2 | 0,05 | 2,46E-02 |
| GO:0051280 | negative regulation of release of sequestered calcium ion into cytosol | 12 | 2 | 0,30 | 2,46E-02 |
| GO:1905116 | positive regulation of lateral attachment of mitotic spindle microtubules to kinetochore | 1 | 1 | 0,02 | 2,46E-02 |
| GO:0060735 | regulation of eIF2 alpha phosphorylation by dsRNA | 1 | 1 | 0,02 | 2,46E-02 |
| GO:2001229 | negative regulation of response to gamma radiation | 1 | 1 | 0,02 | 2,46E-02 |
| GO:0032078 | negative regulation of endodeoxyribonuclease activity | 1 | 1 | 0,02 | 2,46E-02 |
| GO:1903919 | negative regulation of actin filament severing | 1 | 1 | 0,02 | 2,46E-02 |
| GO:0045975 | positive regulation of translation, ncRNA-mediated | 1 | 1 | 0,02 | 2,46E-02 |
| GO:0015964 | diadenosine triphosphate catabolic process | 1 | 1 | 0,02 | 2,46E-02 |
| GO:0033316 | meiotic spindle assembly checkpoint | 1 | 1 | 0,02 | 2,46E-02 |
| GO:0106045 | guanine deglycation, methylglyoxal removal | 1 | 1 | 0,02 | 2,46E-02 |
| GO:0106046 | guanine deglycation, glyoxal removal | 1 | 1 | 0,02 | 2,46E-02 |
| GO:0099606 | microtubule plus-end directed mitotic chromosome migration | 1 | 1 | 0,02 | 2,46E-02 |
| GO:0061360 | optic chiasma development | 1 | 1 | 0,02 | 2,46E-02 |
| GO:0071386 | cellular response to corticosterone stimulus | 1 | 1 | 0,02 | 2,46E-02 |
| GO:0090625 | mRNA cleavage involved in gene silencing by siRNA | 1 | 1 | 0,02 | 2,46E-02 |
| GO:1903096 | protein localization to meiotic spindle midzone | 1 | 1 | 0,02 | 2,46E-02 |
| GO:0051413 | response to cortisone | 1 | 1 | 0,02 | 2,46E-02 |
| GO:1903060 | negative regulation of protein lipidation | 1 | 1 | 0,02 | 2,46E-02 |
| GO:1903073 | negative regulation of death-inducing signaling complex assembly | 1 | 1 | 0,02 | 2,46E-02 |
| GO:1903017 | positive regulation of exo-alpha-sialidase activity | 1 | 1 | 0,02 | 2,46E-02 |
| GO:0007344 | pronuclear fusion | 1 | 1 | 0,02 | 2,46E-02 |

| | | | | | |
|---|---|---|---|---|---|
| GO:0014810 | positive regulation of skeletal muscle contraction by regulation of release of sequestered calcium ion | 1 | 1 | 0,02 | 2,46E-02 |
| GO:1904328 | regulation of myofibroblast contraction | 1 | 1 | 0,02 | 2,46E-02 |
| GO:0036333 | hepatocyte homeostasis | 1 | 1 | 0,02 | 2,46E-02 |
| GO:2000438 | negative regulation of monocyte extravasation | 1 | 1 | 0,02 | 2,46E-02 |
| GO:0032876 | negative regulation of DNA endoreduplication | 1 | 1 | 0,02 | 2,46E-02 |
| GO:0032877 | positive regulation of DNA endoreduplication | 1 | 1 | 0,02 | 2,46E-02 |
| GO:0036471 | cellular response to glyoxal | 1 | 1 | 0,02 | 2,46E-02 |
| GO:1903442 | response to lipoic acid | 1 | 1 | 0,02 | 2,46E-02 |
| GO:1903190 | glyoxal catabolic process | 1 | 1 | 0,02 | 2,46E-02 |
| GO:1903197 | positive regulation of L-dopa biosynthetic process | 1 | 1 | 0,02 | 2,46E-02 |
| GO:1903168 | positive regulation of pyrroline-5-carboxylate reductase activity | 1 | 1 | 0,02 | 2,46E-02 |
| GO:1903181 | positive regulation of dopamine biosynthetic process | 1 | 1 | 0,02 | 2,46E-02 |
| GO:1902483 | cytotoxic T cell pyroptotic process | 1 | 1 | 0,02 | 2,46E-02 |
| GO:1903178 | positive regulation of tyrosine 3-monooxygenase activity | 1 | 1 | 0,02 | 2,46E-02 |
| GO:2000277 | positive regulation of oxidative phosphorylation uncoupler activity | 1 | 1 | 0,02 | 2,46E-02 |
| GO:1903122 | negative regulation of TRAIL-activated apoptotic signaling pathway | 1 | 1 | 0,02 | 2,46E-02 |
| GO:0051692 | cellular oligosaccharide catabolic process | 1 | 1 | 0,02 | 2,46E-02 |
| GO:0051685 | maintenance of ER location | 1 | 1 | 0,02 | 2,46E-02 |
| GO:0090044 | positive regulation of tubulin deacetylation | 1 | 1 | 0,02 | 2,46E-02 |
| GO:0036255 | response to methylamine | 1 | 1 | 0,02 | 2,46E-02 |
| GO:2000775 | histone H3-S10 phosphorylation involved in chromosome condensation | 1 | 1 | 0,02 | 2,46E-02 |
| GO:0021539 | subthalamus development | 1 | 1 | 0,02 | 2,46E-02 |
| GO:1903200 | positive regulation of L-dopa decarboxylase activity | 1 | 1 | 0,02 | 2,46E-02 |
| GO:1901194 | negative regulation of formation of translation preinitiation complex | 1 | 1 | 0,02 | 2,46E-02 |
| GO:0045213 | neurotransmitter receptor metabolic process | 1 | 1 | 0,02 | 2,46E-02 |
| GO:0021633 | optic nerve structural organization | 1 | 1 | 0,02 | 2,46E-02 |
| GO:0009398 | FMN biosynthetic process | 1 | 1 | 0,02 | 2,46E-02 |
| GO:2000597 | positive regulation of optic nerve formation | 1 | 1 | 0,02 | 2,46E-02 |
| GO:2000560 | positive regulation of CD24 production | 1 | 1 | 0,02 | 2,46E-02 |
| GO:0060466 | activation of meiosis involved in egg activation | 1 | 1 | 0,02 | 2,46E-02 |
| GO:0036526 | peptidyl-cysteine deglycation | 1 | 1 | 0,02 | 2,46E-02 |
| GO:0036527 | peptidyl-arginine deglycation | 1 | 1 | 0,02 | 2,46E-02 |
| GO:0036528 | peptidyl-lysine deglycation | 1 | 1 | 0,02 | 2,46E-02 |
| GO:0036529 | protein deglycation, glyoxal removal | 1 | 1 | 0,02 | 2,46E-02 |
| GO:0036530 | protein deglycation, methylglyoxal removal | 1 | 1 | 0,02 | 2,46E-02 |
| GO:0036531 | glutathione deglycation | 1 | 1 | 0,02 | 2,46E-02 |
| GO:0019464 | glycine decarboxylation via glycine cleavage system | 1 | 1 | 0,02 | 2,46E-02 |
| GO:0000461 | endonucleolytic cleavage to generate mature 3'-end of SSU-rRNA from (SSU-rRNA, 5.8S rRNA, LSU-rRNA) | 1 | 1 | 0,02 | 2,46E-02 |
| GO:0046680 | response to DDT | 1 | 1 | 0,02 | 2,46E-02 |
| GO:0035565 | regulation of pronephros size | 1 | 1 | 0,02 | 2,46E-02 |
| GO:0035566 | regulation of metanephros size | 1 | 1 | 0,02 | 2,46E-02 |
| GO:0090144 | mitochondrial nucleoid organization | 1 | 1 | 0,02 | 2,46E-02 |
| GO:0009229 | thiamine diphosphate biosynthetic process | 1 | 1 | 0,02 | 2,46E-02 |
| GO:0009231 | riboflavin biosynthetic process | 1 | 1 | 0,02 | 2,46E-02 |
| GO:1902629 | regulation of mRNA stability involved in cellular response to UV | 1 | 1 | 0,02 | 2,46E-02 |
| GO:0010389 | regulation of G2/M transition of mitotic cell cycle | 182 | 19 | 4,48 | 2,58E-02 |
| GO:0045740 | positive regulation of DNA replication | 36 | 4 | 0,89 | 2,78E-02 |
| GO:0000245 | spliceosomal complex assembly | 64 | 4 | 1,57 | 2,79E-02 |
| GO:0051497 | negative regulation of stress fiber assembly | 27 | 3 | 0,66 | 2,79E-02 |
| GO:0046599 | regulation of centriole replication | 17 | 3 | 0,42 | 2,86E-02 |

| | | | | | |
|---|---|---|---|---|---|
| GO:0000470 | maturation of LSU-rRNA | 26 | 3 | 0,64 | 2,86E-02 |
| GO:1900037 | regulation of cellular response to hypoxia | 11 | 2 | 0,27 | 2,87E-02 |
| GO:0072425 | signal transduction involved in G2 DNA damage checkpoint | 11 | 2 | 0,27 | 2,87E-02 |
| GO:2000786 | positive regulation of autophagosome assembly | 11 | 2 | 0,27 | 2,87E-02 |
| GO:0032968 | positive regulation of transcription elongation from RNA polymerase II promoter | 11 | 2 | 0,27 | 2,87E-02 |
| GO:0061418 | regulation of transcription from RNA polymerase II promoter in response to hypoxia | 70 | 5 | 1,72 | 2,89E-02 |
| GO:1902036 | regulation of hematopoietic stem cell differentiation | 70 | 5 | 1,72 | 2,89E-02 |
| GO:0007080 | mitotic metaphase plate congression | 42 | 6 | 1,03 | 3,02E-02 |
| GO:0000086 | G2/M transition of mitotic cell cycle | 232 | 23 | 5,71 | 3,02E-02 |
| GO:0070498 | interleukin-1-mediated signaling pathway | 96 | 6 | 2,36 | 3,11E-02 |
| GO:0009060 | aerobic respiration | 76 | 9 | 1,87 | 3,26E-02 |
| GO:0051897 | positive regulation of protein kinase B signaling | 151 | 8 | 3,72 | 3,33E-02 |
| GO:0007020 | microtubule nucleation | 29 | 3 | 0,71 | 3,37E-02 |
| GO:0006335 | DNA replication-dependent nucleosome assembly | 29 | 3 | 0,71 | 3,37E-02 |
| GO:1903214 | regulation of protein targeting to mitochondrion | 41 | 3 | 1,01 | 3,38E-02 |
| GO:0007077 | mitotic nuclear envelope disassembly | 12 | 2 | 0,30 | 3,39E-02 |
| GO:0071712 | ER-associated misfolded protein catabolic process | 12 | 2 | 0,30 | 3,39E-02 |
| GO:0016446 | somatic hypermutation of immunoglobulin genes | 12 | 2 | 0,30 | 3,39E-02 |
| GO:0031936 | negative regulation of chromatin silencing | 12 | 2 | 0,30 | 3,39E-02 |
| GO:0060213 | positive regulation of nuclear-transcribed mRNA poly(A) tail shortening | 12 | 2 | 0,30 | 3,39E-02 |
| GO:0006521 | regulation of cellular amino acid metabolic process | 52 | 5 | 1,28 | 3,40E-02 |
| GO:0097421 | liver regeneration | 30 | 3 | 0,74 | 3,68E-02 |
| GO:0033169 | histone H3-K9 demethylation | 13 | 2 | 0,32 | 3,94E-02 |
| GO:0070365 | hepatocyte differentiation | 13 | 2 | 0,32 | 3,94E-02 |
| GO:0006298 | mismatch repair | 31 | 3 | 0,76 | 4,00E-02 |
| GO:0006271 | DNA strand elongation involved in DNA replication | 18 | 3 | 0,44 | 4,50E-02 |
| GO:0007076 | mitotic chromosome condensation | 14 | 2 | 0,34 | 4,52E-02 |
| GO:0045787 | positive regulation of cell cycle | 339 | 26 | 8,34 | 4,54E-02 |
| GO:0030097 | hemopoiesis | 802 | 30 | 19,74 | 4,57E-02 |
| GO:0001701 | in utero embryonic development | 311 | 11 | 7,65 | 4,62E-02 |
| GO:0006296 | nucleotide-excision repair, DNA incision, 5'-to lesion | 33 | 3 | 0,81 | 4,68E-02 |
| GO:0045841 | negative regulation of mitotic metaphase/anaphase transition | 36 | 9 | 0,89 | 4,77E-02 |

***Supplementary Table S6: All 11 significant HLA alleles divided per population group related to development of HZ in patients versus controls*. *=statistical significance after multiple testing correction.**

| | **Asian** | **Black** | **White** |
|---|---|---|---|
| **HLA allele** | **Odds ratio (± 95% CI)** | **Odds ratio (± 95% CI)** | **Odds ratio (± 95% CI)** |
| **A*01:01** | 0,698 (0,326 – 1,365) | 2,553 (0,752 – 6,927) | 1,162* (1,076 – 1,255) |
| **B*07:02** | 0,470 (0,055 – 1,794) | 1,485 (0,438 – 4,030) | 1,158* (1,067 – 1,257) |
| **C*07:02** | 0,902 (0,464 – 1,661) | 1,218 (0,305 – 3,578) | 1,161* (1,071 – 1,257) |
| **B*40:01** | 0,671 (0,134 – 2,088) | 0,000 (0,000 – 14,011) | 1,229* (1,094 – 1,377) |
| **A*33:03** | 0,532 (0,203 – 1,183) | 0,258 (0,006 – 1,573) | 0,397 (0,108 – 1,024) |
| **DRB3*02:02** | 0,757 (0,440 – 1,287) | 0,384 (0,138 – 0,935) | 0,884 (0,809 – 0,966) |
| **B*44:02** | 1,633 (0,190 – 6,366) | 0,000 (0,000 – 7,151) | 0,812* (0,735 – 0,896) |
| **DRB1*11:01** | 0,887 (0,310 – 2,082) | 0,385 (0,044 – 1,547) | 0,776 (0,654 – 0,915) |
| **A*02:01** | 0,807 (0,282 – 1,891) | 0,831 (0,246 – 2,236) | 0,844* (0,783 – 0,910) |
| **DQB1*03:01** | 1,041 (0,560 – 1,862) | 1,165 (0,478 – 2,675) | 0,860* (0,791 – 0,933) |
| **C*05:01** | 0,807 (0,020 – 4,808) | 0,000 (0,000 – 3,173) | 0,834* (0,755 – 0,919) |
